# Global practices in paediatric olfactory dysfunction: a cross-sectional survey of paediatric ENT surgeons

**DOI:** 10.64898/2026.06.04.26354942

**Authors:** Gina M. Spencer, Kunwar Karim, Agnieszka Dzioba, M. Elise Graham, Peng You, Thomas Hummel, Janine Gellrich, Paula Coyle, Hannah Burns, Shazia Peer, Faisal Zawawi, Jérome R. Lechien, Valentin A. Schriever, Eishaan K. Bhargava, Katherine L. Whitcroft

## Abstract

**Background:** Olfactory dysfunction (OD) in children remains underdiagnosed and poorly characterised. Despite its known impacts on nutrition, quality of life, safety awareness, and psychosocial development, no standardised diagnostic or management pathway currently exists for paediatric OD. This study aimed to characterise global practice patterns and identify diagnostic and therapeutic challenges unique to paediatric care.

**Methodology/Principal:** A 44-item cross-sectional online survey was distributed to a verified international network of paediatric otolaryngologists across 36 countries via a closed professional platform. The survey assessed five domains: diagnostic practices, management protocols, technology and innovation, education and training, and barriers to effective care. Regional grouping was used to facilitate meaningful statistical comparisons. Categorical variables were evaluated using chi-square tests, with odds ratios and 95% confidence intervals reported for significant findings.

**Results:** Of 351 potential participants, 167 responded (47.6% response rate). Most respondents (83%) reported seeing children with OD, yet 95% saw fewer than ten such patients annually. Psychophysical testing was never performed by 54.8% of respondents, while 88.4% routinely ordered cross-sectional imaging. Testing frequency increased significantly with patient age (Cochran’s Q p<0.001). The most common barriers to objective testing were insufficient training (44.3%), time constraints (29.9%), and funding limitations (28.1%). Multidisciplinary collaboration was negligible. Significant regional variation was observed across most practice domains.

**Conclusions:** Paediatric OD care is characterised by functional underinvestigation, fragmented multidisciplinary collaboration, and systemic educational gaps. These findings support urgent development of standardised clinical guidelines, age-appropriate validated assessment tools, and formal interdisciplinary care pathways.

## INTRODUCTION

Olfactory dysfunction (OD) has been reported to affect ∼22% of the adult population.(^1^) OD in children, however, remains underdiagnosed and poorly characterised, with a paucity of epidemiological studies exploring this issue that can significantly impact nutritional intake and quality of life (QoL).(^2–4^) While underexplored in children, adults often report reduced food enjoyment, which can lead to eating disorders and depression.(^5^) There is also a need to address safety concerns in OD, like detecting smoke, gas leaks, or spoiled food.(^6^) In studies that do attempt to classify the epidemiology of OD, diagnosis of the dysfunction is often based on self-report, with standardized olfactory testing only occurring in a minority.(^7,8^) A recent scoping review has called for all clinicians to have a diagnostic algorithm for identifying the cause of OD when they encounter it in practice, given the diverse range of aetiologies and potential treatment methods.(^9^)

There are numerous challenges in assessing olfactory function in paediatric populations, including the use of odour stimuli that may be unfamiliar to children and the cognitive and linguistic demands required by many existing tests.(^10^) Tools such as threshold detection and odour identification have been adapted for use in children.(^11^) However, even in adult populations, olfactory testing is often limited by economic, educational, and structural barriers, including funding constraints, time pressures, inadequate training, staffing shortages, and a lack of recognition of smell testing as standard care.

A recent international survey of practice highlighted gaps and variations in olfactory assessment practices among ENT clinicians.(^12^) This study serves as the paediatric counterpart to that adult benchmark, aiming to characterise global practice patterns and the specific diagnostic and therapeutic challenges unique to paediatric care.

## MATERIALS AND METHODS

### Questionnaire development

Ethics approval for this cross-sectional survey was obtained from the Health Sciences Research Ethics Board at Western University (REB # 126849). The survey questions were informed by clinical practice guidelines and existing literature identifying gaps in current practice.(^12^) An expert panel comprising international topical experts in olfaction (KW, TH, JG) and paediatric otolaryngologists from nine countries contributed to questionnaire development and refinement, ensuring content validity. Face validity was assessed through qualitative feedback during questionnaire piloting with experts and target group members.

The 44-item anonymous, online survey presented questions in multiple formats, including multiple choice, select multiple options, and open-text fields. It assessed demographic information, along with five domains surrounding specifics of paediatric OD practice patterns: diagnostic practices, management protocols, technology and innovation in practice, education and training, and challenges to effective care.

The survey was available to be completed digitally for six weeks from May 16 to June 27, 2025. Two reminders were sent at two-week intervals. The survey was hosted at London Health Sciences Centre Research Institute (London, Ontario, Canada) using REDCap electronic data capture, a secure and encrypted online platform.(^13,14^) Participation was voluntary. The complete survey is included as Supporting Information.

### Participant selection

Participants were invited from a closed, verified professional network of paediatric otolaryngologists via an international WhatsApp group. A maximum of 351 potential participants were identified. The WhatsApp group administrators confirm the identity of all members as practicing paediatric otolaryngologists prior to admission to the group. English proficiency and current practice as a paediatric otolaryngologist were the inclusion criteria. Participants were recruited from 36 countries, offering a diverse and international pool for survey distribution. An accurate response rate calculation was possible through direct recruitment on this platform.

### Statistical analysis

Regional grouping of participants was employed to mitigate the impact of small sample sizes in individual countries, facilitate meaningful statistical comparisons, and align with approaches used in prior international physician surveys which similarly grouped countries to enable analysis of global patterns while maintaining interpretability.(^15,16^) Descriptive statistics, including frequencies, percentages, medians, and interquartile ranges (IQR), were used to summarize respondent characteristics and key survey variables. Categorical variables were evaluated using chi-square tests. Participants with incomplete survey responses (n=33) were included in analyses where applicable, with missing data excluded on a per-question basis. Statistical significance was defined as *p*<0.05. For statistically significant findings, odds ratios (OR) with corresponding 95% confidence intervals (CI) are presented. All data analyses were performed using SPSS Statistics version 29 (IBM Corp., Armonk, NY, USA).

## RESULTS

### Participant demographics

Responses meeting inclusion criteria were received from 167 of 351 potential participants identified, yielding a 47.6% response rate. 134 of these individuals completed the entire survey (80.2% of total respondents). 100% of respondents provided their country of practice. These countries were grouped into the following regions for data analysis: Africa (n=5, 3.0%), Asia (n=16, 9.6%), Australia (n=27, 16.2%), Europe (n=23, 13.8%), Middle East (n = 18, 10.8%), Canada/Carribean (n=10, 6.0%), South America (n=7, 4.2%), United Kingdom (n=39, 23.4%), United States of America (n=22, 13.2%). The geographic distribution of respondents is displayed in Figure 1.

**Figure 1.**
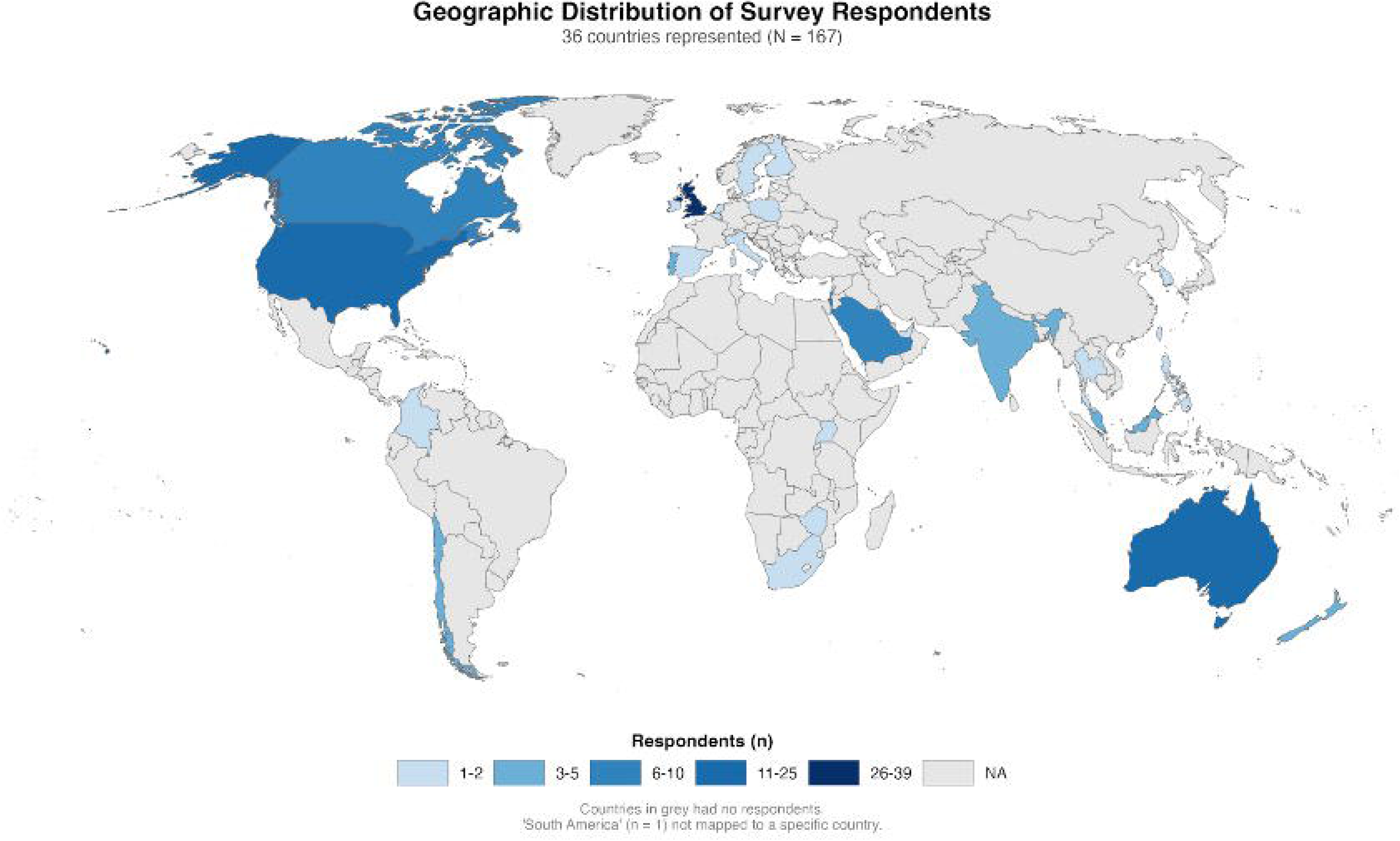
Geographic distribution of survey respondents by country. Thirty-six countries were identified amongst our respondents, with most from United Kingdom (n=39, 23.4%), followed by Australia (n=22, 13.2%) and the USA (n=22, 13.2%). Additional representation included Canada/Caribbean (n=10, 5.4%), France (n=8, 4.8%), Israel (n=6, 3.6%), Saudi Arabia (n=6, 3.6%), India (n=5, 3.0%), New Zealand (n=5, 3.0%), Chile (n=4, 2.4%), Malaysia (n=3, 1.8%), and Portugal (n=3, 1.8%).

Distribution of respondent characteristics by geographic region is depicted in Supplementary Figure 1, including years in practice and subspecialty interests. Significant regional variation was observed (χ² p<0.001). General paediatric ENT (defined as respondents who practice a full scope of paediatric ENT without further subspecialty) (76%, n=127) was the predominant subspecialty across all regions, with statistically significant regional variation in general ENT (5.4%, n=9) and rhinology (19.8%, n=33) representation.

### Current assessment practices

Eighty-three percent of respondents reported seeing children with OD, though the majority (61.3%) encountered these cases less than monthly (p=.427) (Figure 2). General paediatric ENT clinics were the predominant assessment setting (71.3%), with significant regional variation in private clinic (22.8%, p<.001) and specialist rhinology clinic use (10.8%, p=.050) (Supplementary Figure 2).

**Figure 2.**
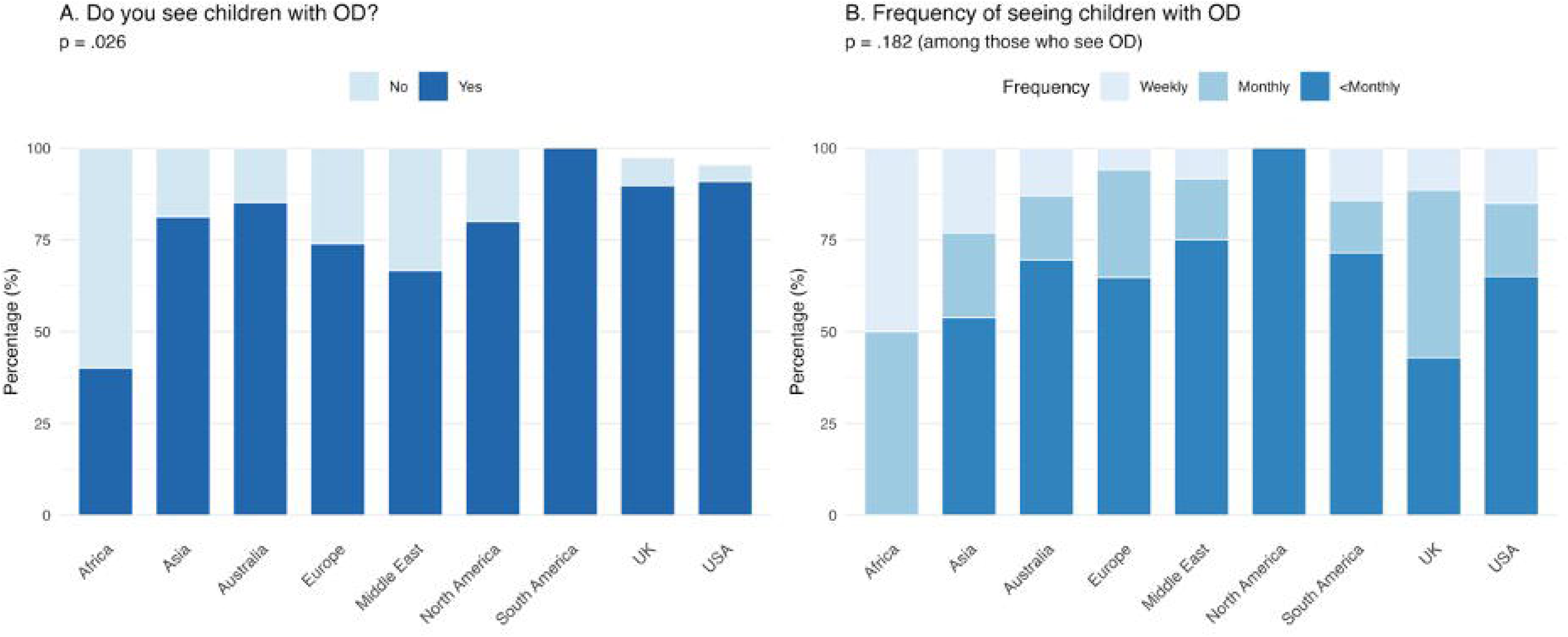
Current assessment practices for paediatric OD by region. (A) Proportion of respondents reporting that they see children with OD in their clinical practice (83% overall, *p*=.026). (B) Frequency of encounters among those who see OD. Percentages calculated relative to regional respondent totals for Panel A, and relative to those who see OD for Panels B-D. Regional sample sizes: Africa n=5, Asia n=16, Australia n=27, Europe n=23, Middle East n=18, North America n=10, South America n=7, UK n=39, USA n=22.

Primary care providers (49.1%, p<.001) and general paediatricians (40.1%, p=.330) were the most frequent referral sources, with parental self-referral (24.6%) particularly common in the UK and Australia. Once anosmia was identified, majority of respondents (81.8%, n=108/132) elected to manage patients within their own practice.

Most clinicians (54.8%) reported never performing psychophysical tests, while routine testing (>75% of cases) was reported by 15.3% of respondents, predominantly from the USA, Europe, and UK. Testing frequency increased significantly with age (Cochran’s Q p<.001): testing was minimal in children aged 0–2 years (0.6%) and 3–6 years (8.4%), rising in those aged 7–12 years (26.3%) and >12 years (28.1%). Europe, UK, and USA were significantly more likely to test older children (7–12 years: p=.019; >12 years: p=.010) (Figure 3).

**Figure 3.**
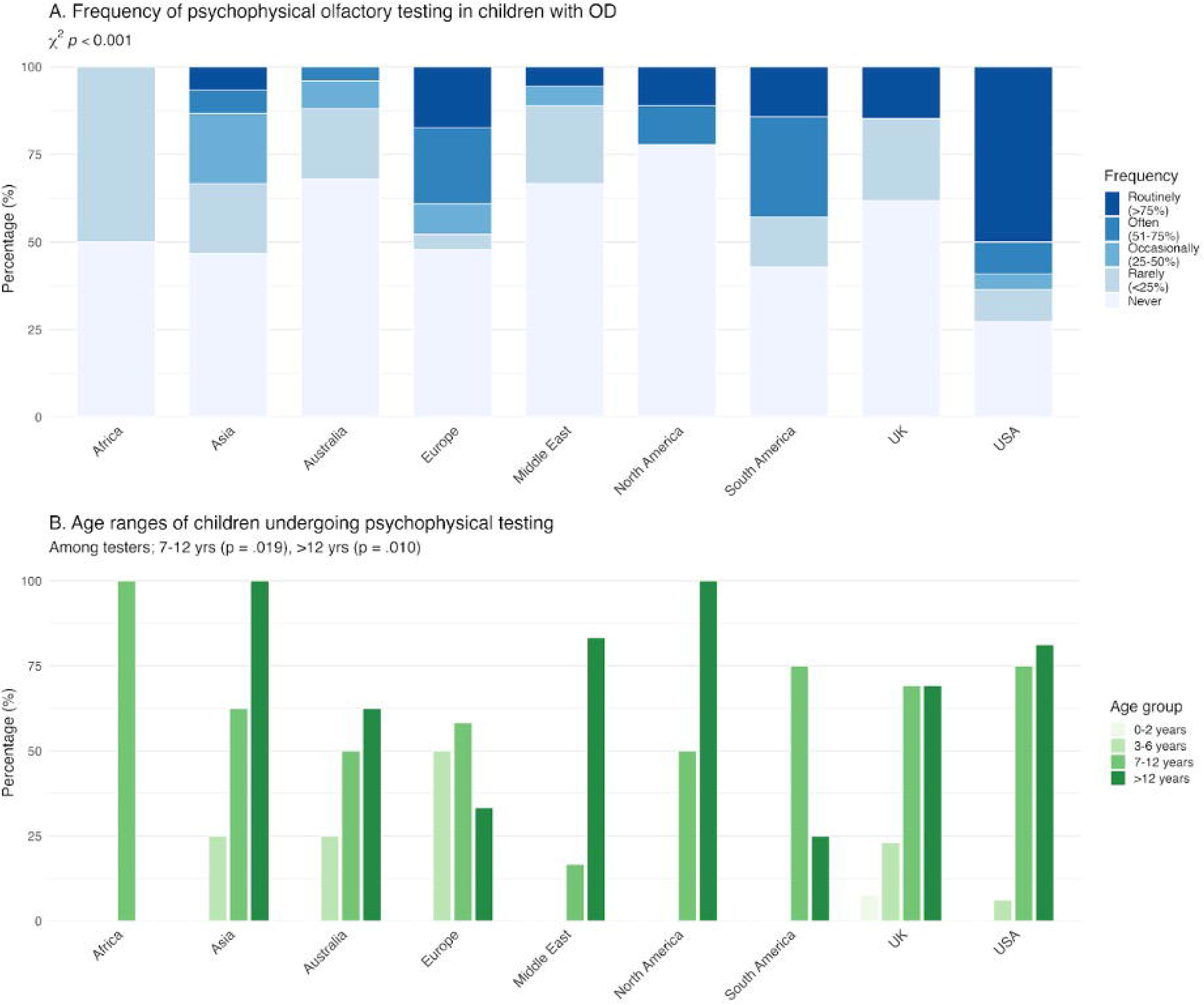
Psychophysical olfactory testing practices in children with OD by region. (A) Frequency of psychophysical testing across all respondents who see children with OD, showing significant regional variation (χ² *p*<.001). (B) Age ranges of children undergoing psychophysical testing, among those who perform such assessments. Percentages in Panel A are calculated relative to all regional respondents; percentages in Panel B are relative to those who perform testing in each region. Regional sample sizes: Africa n=5, Asia n=16, Australia n=27, Europe n=23, Middle East n=18, North America n=10, South America n=7, UK n=39, USA n=22.

Validated tool use remained uncommon across all age groups and showed strong age-dependence, with clinicians in the youngest age bands relying primarily on parental report and behavioural observation. Standardised tools became more prevalent from age 7 onwards, though significant regional variation was observed for the Paediatric Smell Wheel (p=.008) and University of Pennsylvania Smell Identification Test (UPSIT) without paediatric norms (p=.007). In the >12-year group, adapted adult protocols predominated (19.8%, p=.015), with highest use in the USA (50%). Full tool-use data stratified by age group and region are presented in Supplementary Table 1.

Key barriers to objective smell testing are presented in Figure 4 and Supplementary Table 2. Insufficient experience or training in olfactory assessment was the most commonly cited barrier (n=74, 44.3%), followed by insufficient time (n=50, 29.9%) and institutional funding limitations (n=47, 28.1%).

**Figure 4.**
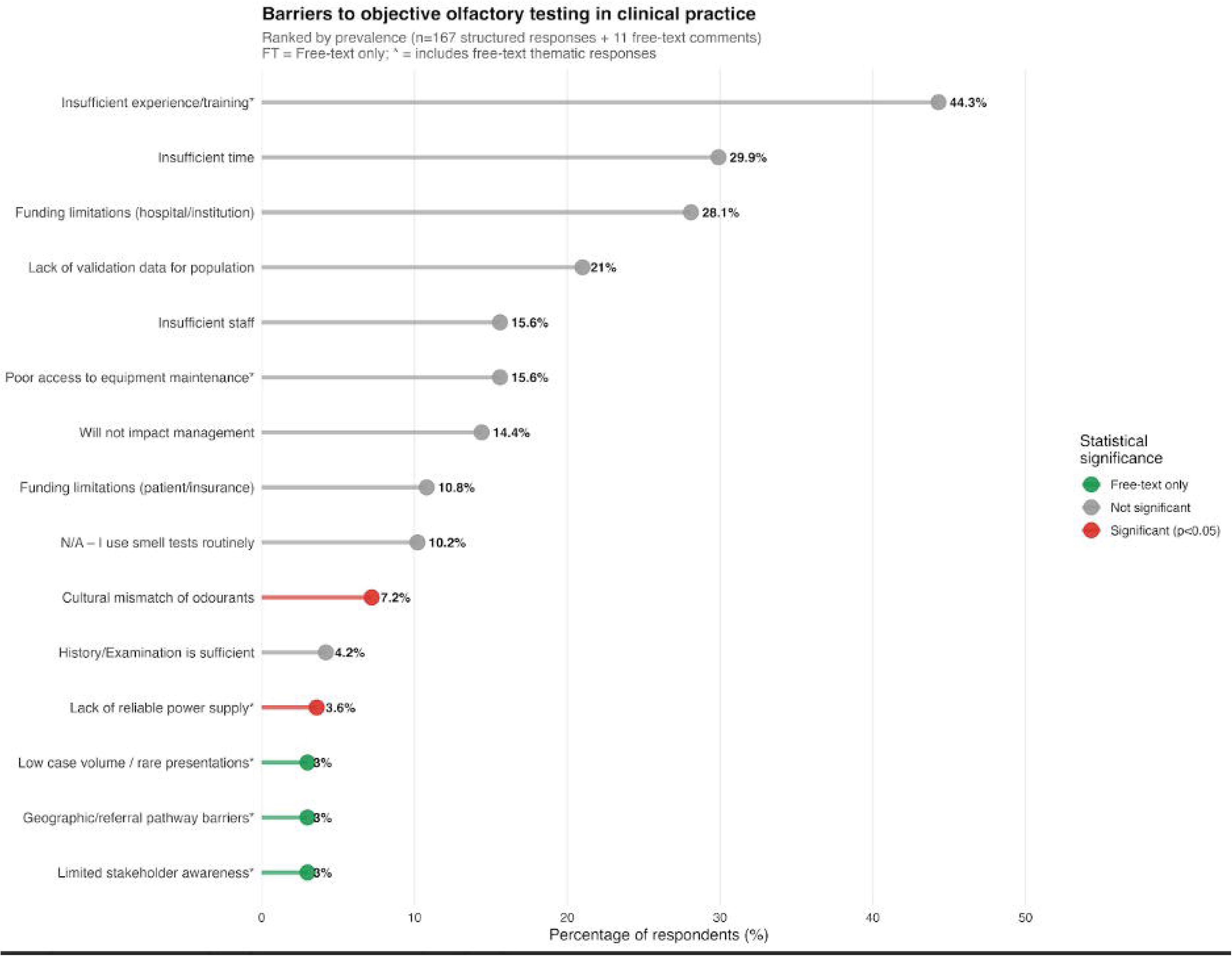
Barriers to objective olfactory testing in clinical practice, ranked by prevalence. Lollipop chart showing the percentage of respondents reporting each barrier to implementing psychophysical smell tests in children with OD. Ranked prevalence of barriers combining structured survey responses (n=167) and analysis of free-text responses (n=11 respondents). Barriers are ordered from most to least frequently cited among structured responses. Point colour indicates: red = statistically significant regional variation (*p*<0.05); orange = marginal significance (*p*<0.10); grey = no significant regional variation; green = identified exclusively through free-text responses. An asterisk (*) indicates barriers with substantive free-text elaboration or clarification. Labels show percentage of respondents; “FT” indicates free-text-derived themes without quantitative prevalence data. Regional sample sizes: Africa n=5, Asia n=16, Australia n=27, Europe n=23, Middle East n=18, North America n=10, South America n=7, UK n=39, USA n=22.

### Management approaches

Overall, 88.4%The vast majority of respondents (88.4%, n=137/155) reported they would organize cross-sectional imaging when OD is the presenting or isolated symptom, with no significant regional variation (p=.065) (Supplementary Figure 3).

Proactive screening was most frequently reported for post-viral OD (43.4%) and Kallmann syndrome/hypogonadotropic hypogonadism (35.9%); rates fell below 15% for most syndromic conditions, including CHARGE syndrome (11.0%, p=.043), eating disorders (9.7%, p=.003), and autism spectrum disorder (6.0%, p=.043) (Figure 5).

**Figure 5.**
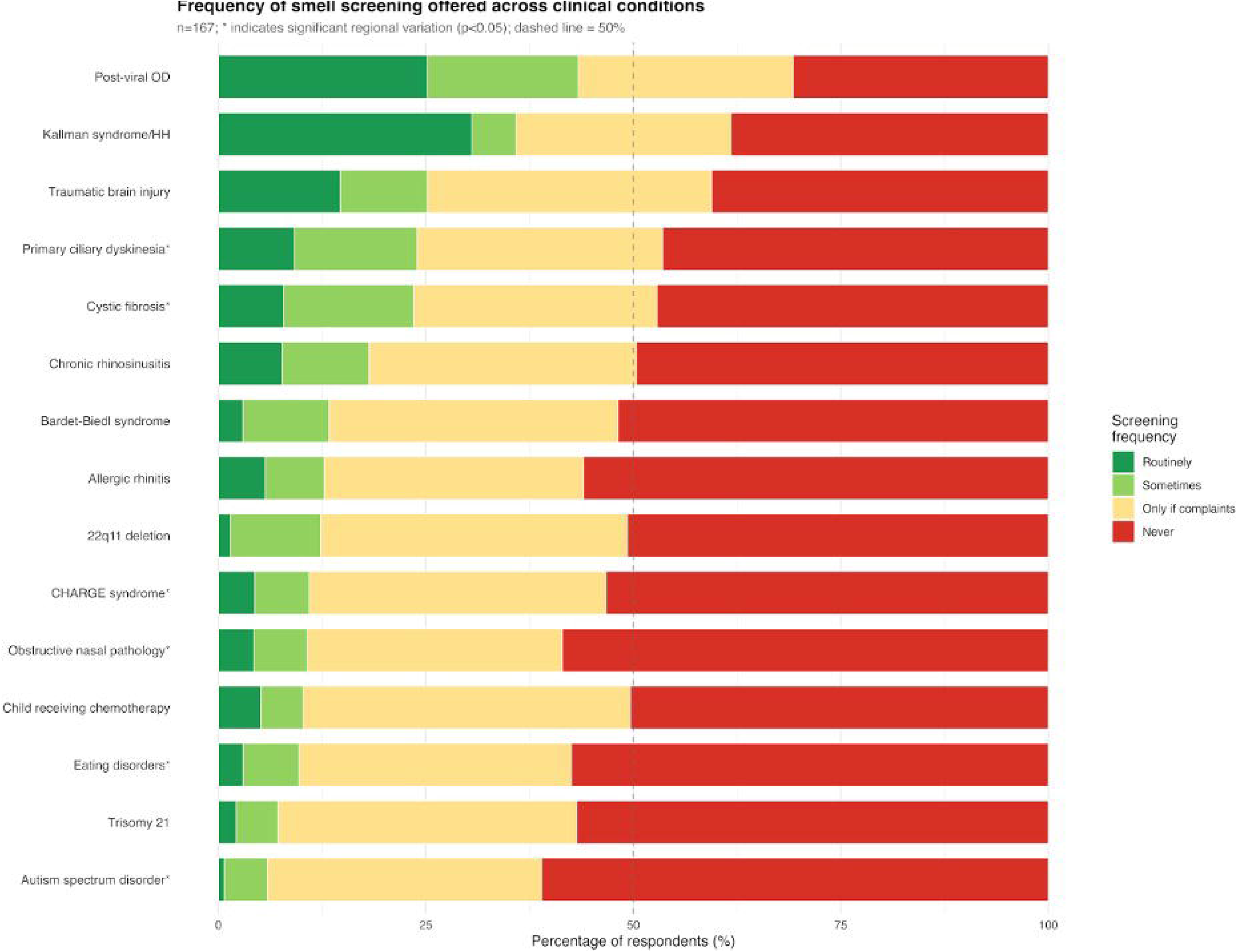
Frequency of proactive smell screening offered across clinical conditions: Stacked bar chart shows the proportion of respondents who would offer smell screening routinely (dark green), sometimes (light green), only if the child complains of olfactory dysfunction (yellow), or never (red) across 15 clinical conditions (n=167 respondents). Conditions are ranked by the combined proportion of proactive screening (routinely + sometimes). Asterisk (*) indicates significant regional variation (χ² p<0.05). Dashed vertical line indicates 50%. Regional sample sizes: Africa n=5, Asia n=16, Australia n=27, Europe n=23, Middle East n=18, North America n=10, South America n=7, UK n=39, USA n=22.

Management protocols for congenital anosmia are displayed in Figure 6; full regional breakdowns are provided in Supplementary Table 3. Endocrine referral (50.9%) and full neurological examination (49.7%) were the most consistently adopted assessments across regions. Genetic counselling showed the greatest regional divergence (29.9% overall, p<.001), with high uptake in the USA (64%), Europe (44%), and South America (43%) contrasting with minimal adoption in Australia (7%) and UK (18%). Safety counselling was offered by 38.9% of respondents overall, with the highest rates in the USA (55%) and South America (43%); peer support referral remained rare globally (4.8%).

**Figure 6.**
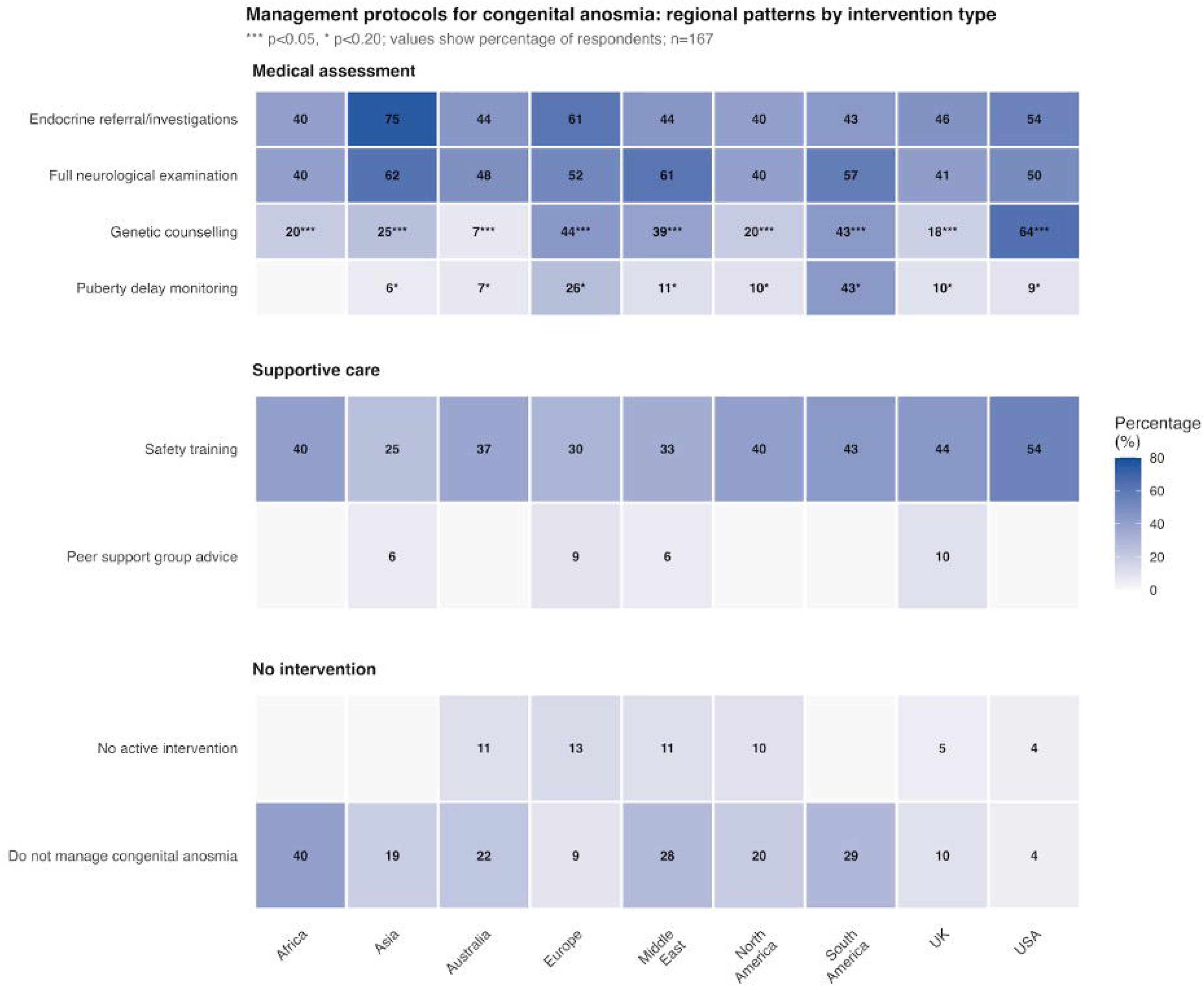
Management protocols for congenital anosmia stratified by intervention type and region: Heatmap showing the percentage of respondents who would include each management component in their protocol for congenital anosmia, grouped by intervention type: medical assessment (top panel), supportive care (middle panel), and no intervention (bottom panel). Cell colour intensity indicates adoption rate, with values displayed as percentages. Asterisks denote statistical significance of regional variation: *** indicates *p*<0.05, * indicates *p*<0.20. Regional sample sizes: Africa n=5, Asia n=16, Australia n=27, Europe n=23, Middle East n=18, North America n=10, South America n=7, UK n=39, USA n=22.

Olfactory training (OT) was the most commonly adopted intervention for chronic OD (35.9%), with highest uptake in South America (71%), USA (46%), and Middle East (44%) (Supplementary Figure 4. Among the minority performing repeat psychophysical testing (n = 24), annual follow-up predominated (45.8%), followed by bi-annual (41.7%) and quarterly (12.5%) schedules (p=.086). Free-text responses highlighted the absence of formal follow-up infrastructure in many settings: “No defined clinic for this” (Canada/Caribbean) and “Variable. No protocol” (USA) were representative comments.

Multidisciplinary collaboration was inconsistent and highly variable by region (Supplementary Figure 5; Supplementary Table 4). Despite eating difficulties being the most frequently recognised QoL impact (47.3%), involvement of dietitians (1.7% routinely in syndromic OD), speech and language pathology (5.8%), and neuropsychology (2.6%) was negligible. Lack of referral guidelines was the most cited barrier to collaboration (41.9%), with lack of shared care pathways (34.7%) and fragmented healthcare systems (22.8%) also frequently reported (Supplementary Figure 6). Institutional funding limitations showed marked regional variation (18.6% overall; range: 0–60%, p<.001).

### Barriers to effective care

Inadequacy of training in diagnosis and management of olfactory disorders (50.3%, n=84), high cost of objective smell tests (38.9%, n=65), and inadequately trained ancillary staff (37.7%, n=63) were the top identified barriers to effective care of patients with OD. 10.8% (n=18) reported a lack of availability of culturally appropriate odourants for objective testing. Free text responses included “lack of awareness to this entity,” “patients do not feel the importance,” and “[healthcare decision makers/administrators] need to channel scarce resources into more pressing areas.”

Lastly, respondents identified top advocacy priorities for paediatric OD. These included better olfactory training during residency (44.3% of respondents, n=74), integration of olfactory assessment/screening in primary care (32.9%, n=55), and national smell screening guidelines (31.7%, n=53). Additionally, 20.4% of respondents (n=34) selected institution/public funding for smell rehabilitation, and 10.8% (n=18) selected insurance coverage. School accommodations for anosmic children were considered important by 7.8% (n=13). Meanwhile, 7.8% (n=13) did not feel that OD requires advocacy in the field today.

## DISCUSSION

Our cross-sectional survey study sought to better understand the international practice variations in diagnosis and management of OD in the paediatric population, with a paucity of data currently reported in scientific literature.(^9^) We acknowledge that clinical practices vary by region and are often shaped by national or international guidelines; clinicians are therefore encouraged to follow the recommendations relevant to their respective jurisdictions.

### Age-related assessment gaps and clinical implications

Despite an estimated adult OD prevalence of approximately 22%, the vast majority of respondents (95%) reported seeing fewer than 10 paediatric patients per year. Similarly, only a small proportion of patients seen at specialized centers for smell and taste disorders—around 2 to 4%—are children or adolescents.(^11^) This disparity that may reflect underdiagnosis, under-referral, or a genuinely lower burden of disease in younger populations.(^1^) Unawareness of olfactory loss is well-documented in adults,(^17^) partly because olfaction operates largely outside of conscious perception.(^18^) This phenomenon is likely to be amplified in children, who often lack the vocabulary, insight, or comparative life experience to recognize and articulate changes in their sense of smell. Caregivers may similarly normalise the symptom in the absence of overt consequences such as safety incidents or nutritional decline.

The long-term developmental impact of OD, whilst understudied, is not trivial. Isolated congenital anosmia (ICA) is associated with increased social insecurity, a greater risk of depressive symptoms, and a higher incidence of household accidents.(^19^) It is also associated with significantly fewer sexual relationships in men and lower perceived relationship security in women.(^20^) These findings reinforce that the low clinical encounter rate likely reflect a detection gap rather than a benign disease burden.

Testing frequency increased significantly with age in our cohort (Cochran’s Q p<.001), consistent with the known developmental trajectory of olfactory performance - driven by olfactory system maturation, broadening odour exposure, and expanding vocabulary.(^11,21^) Even with paediatric-adapted tools such as Sniffin’ Sticks,(^21^) the paediatric Smell Wheel,(^22^) and the paediatric Barcelona Olfactory Test (pBOT)(^23,24^) and the Universal Sniff (U-Sniff)(^25^) (the only test developed de novo for children with multinational normative data) performance remains susceptible to confounding by cognitive and linguistic development, making it challenging to isolate true olfactory deficits from age-related perceptual and cognitive limitations.(^11,26^) In practice, with younger age groups, clinicians must therefore rely on caregiver report, behavioural observation, and adjunctive imaging, accepting the inherent limitations of these approaches.

### Barriers to olfactory testing

Barriers to testing were dominated by insufficient training (44.3%) and time constraints (29.9%), suggests that educational initiatives and workflow optimization may be critical targets for intervention. Infrastructure barriers such as funding limitations (28.1%), inadequate staff (15.6%), and lack of access to equipment maintenance and reliable power supply (25.6% and 3.6% respectively) likely highlight disparities in resource availability across regions.

The perceived lack of validation data in pediatric populations (21%) highlights a critical gap in the literature, as many existing psychophysical smell tests have been developed and validated primarily in adult cohorts, limiting clinician confidence in their applicability to children.

Notably, 14.4% of respondents believed that test results would not meaningfully influence management, a perception that sits at odds with growing evidence supporting OT and targeted aetiological treatment as effective interventions for paediatric OD. Addressing this belief through targeted education and dissemination of outcome data is perhaps as important as developing the tools themselves.

### Assessment practice variations

The high overall uptake of imaging (88.4%) contrasts sharply with low rates of psychophysical testing (54.8% never test), suggesting that structural investigations are prioritized over functional assessment in current practice. The predominance of MRI protocols targeting olfactory bulb anatomy (47.3%) and neoplasm exclusion (41.9%) perhaps reflects clinical concern for congenital anomalies and mass lesions rather than functional characterization. Significant regional variation in imaging strategies indicates heterogeneity in diagnostic algorithms, while geographically concentrated funding barriers highlight access inequities in resource-limited settings.

### Management approaches beyond watchful wait

The literature does not support a single, standardized treatment for paediatric OD, reflecting the heterogeneity of its underlying causes.(^9^) Few studies to date have examined the benefit of OT for OD in the paediatric population.(^9^) One study reported improvement in odour identification ability, but not olfactory thresholds, with six weeks of OT in normosmic 8-year-old children.(^27^) Another study in children ages 6-16 years with OD after mild traumatic brain injury (mTBI) reported accelerated rehabilitation of olfactory sensitivity with OT using low-concentration odours.(^28^)

Despite emerging evidence supporting its effectiveness, less than half of our respondents reported offering OT to patients with chronic OD (35.9%) and acute post-infectious olfactory dysfunction (PIOD) (45.5%), with significant variation by geographic region (*p*=.014) that may reflect differences in access to OT resources, clinician familiarity with evidence-based protocols, or varying local guidelines. The relatively low global uptake, even in regions with higher response rates, suggests a need for increased awareness and implementation of OT as a first-line, low-risk intervention for acquired OD.

In children with chronic OD, only 12.6% included dietary or nutritional assessments in their management. This is concerning given emerging evidence linking olfactory deficits with feeding challenges in children.(^29–31^) A recent study of 246 children aged 3 to 9 demonstrated that poorer odour identification and reduced sensitivity to non-food odours were all significant predictors of food neophobia.(^32^) These findings highlight olfaction’s role in early eating behaviors. The low dietary assessment and dietitian involvement indicate a care gap, risking missed early interventions for children with potential nutritional or feeding issues due to OD.

### The multidisciplinary care deficit

Hummel et al., in alignment with the current guideline from the Association of the Scientific Medical Societies in Germany, recommend that suspected congenital OD based on clinical or imaging findings should prompt interdisciplinary evaluation involving paediatricians, endocrinologists, and, where possible, geneticists.(^33^) Despite these recommendations, in practice, the diagnostic and management pathway for paediatric OD is often fragmented, lengthy, and characterized by multiple referrals across specialties, if at all.(^23^)

Our survey findings reinforce this reality, with interdisciplinary collaboration in the management of children with OD remaining inconsistent and highly variable by region. Engagement with dieticians was particularly poor, with only 1.7% of respondents reporting routinely involving dietitians in children with syndromic OD, despite the clear relevance of dietary support in cases where OD contributes to selective eating, nutritional deficiencies, or further feeding challenges in the context of a complex clinical presentation. This parallels low engagement with paediatric neurology (15.5%), clinical genetics (21.4%), speech-language pathology (5.8%), and neuropsychology (2.6%) - all of whom play arguably essential roles in the comprehensive assessment of syndromic and complex paediatric patients. The absence of significant variation in referral guidelines barriers (*p*=0.390) despite marked differences in collaboration rates indicates that lack of guidelines is a widespread, systematic problem affecting most regions rather than a localized issue. Guideline development and dissemination may be a high-impact intervention to improve multidisciplinary care coordination for paediatric OD.

### Strengths and limitations

As this is a survey-based study, our findings are subject to inherent selection and response biases. The use of convenience sampling through professional networks reduces generalizability; however, it is a commonly used approach in international physician surveys and has been shown to effectively capture perspectives from clinicians actively engaged in the field.(^34–39^) Certain regions - particularly Africa, South America, and parts of Asia - were underrepresented. This imbalance could further influence the generalizability of our findings and the statistical power to detect regional differences. To partially mitigate this, countries were grouped into broader regions (e.g., Europe, North America, Middle East) following approaches previously reported in international physician surveys, which allowed for more meaningful regional comparisons while maintaining consistency with prior literature.(^15,16^) Despite these steps, the uneven regional distribution underscores the need for future efforts to engage a more globally representative cohort, ideally through multi-lingual survey instruments.

Nonetheless, by establishing the first global baseline of paediatric OD practice across 36 countries, with a response rate of over 45%, this study provides the evidence needed to drive the next critical steps: standardised clinical guidelines, targeted educational initiatives, and the interdisciplinary care pathways that children with OD currently lack.

### Future directions and clinical implications

This survey identifies five priority areas to advance paediatric OD care globally:

1. Awareness and Education – Public awareness programs are needed to increase awareness of pediatric OD, its early signs, and potential impacts on development and QoL. Targeted educational initiatives for pediatricians and otolaryngologists are needed to improve early recognition, given the low rates of proactive olfactory screening reported across regions.
2. Screening and Early Identification – Routine olfactory assessment strategies should be developed for at-risk groups, including children with congenital OD, post-viral OD, and feeding or sensory integration difficulties.
3. Interdisciplinary Collaboration – Formal care pathways incorporating dietitians, clinical geneticists and neuropsychologists are needed, particularly for syndromic presentations and young children unable to complete standard psychophysical tests, to ensure comprehensive evaluation and management.
4. Validated Paediatric Assessment Tools – Investment in developing and disseminating age-appropriate, culturally adapted olfactory tests with normative data across developmental stages alongside olfaction-specific QoL assessments, including caregiver-proxy versions for younger ages, to capture the unique developmental and psychosocial impact of OD remains a critical unmet need.
5. Guideline development – The absence of consensus clinical guidelines is a systemic barrier across all regions; international collaborative efforts should prioritise standardised diagnostic and management frameworks for paediatric OD.

## Conclusion

There is currently no standardized pathway or consensus recommendation for the management of paediatric OD. Our international survey highlights significant global variability in prevalence estimation, diagnostic approaches, interdisciplinary collaboration, follow-up practices, and treatment strategies. Despite this variation, consistent barriers were identified across regions, particularly related to limitations in objective testing and challenges in diagnosing OD in early childhood. These findings should guide the development of standardized clinical practice guidelines and promote the creation of age-appropriate, population-specific diagnostic tools. Ultimately, these steps are essential to improving early detection, intervention, and QoL for children with OD.

## Data Availability

All data produced in the present study are available upon reasonable request to the authors.

## ACKNOWLEDGEMENTS

None

## AUTHORSHIP CONTRIBUTION

Please describe the individual contributions of each author to the paper.

## CONFLICT OF INTEREST

No conflicts of interest to declare.

## FUNDING

None

---

Supplementary Figure 1

Supplementary Figure 2

Supplementary Figure 3

Supplementary Figure 4

Supplementary Figure 5

Supplementary Figure 6

Supplementary Figure 7

Supplementary Figure 8

Supplementary Tables

## Notes

### Competing Interest Statement

The authors have declared no competing interest.

### Author Declarations

Ethics approval for this cross-sectional survey was obtained from the Health Sciences Research Ethics Board at Western University (REB # 126849).

## REFERENCES

1. Desiato VM, Levy DA, Byun YJ, Nguyen SA, Soler ZM, Schlosser RJ. The Prevalence of Olfactory Dysfunction in the General Population: A Systematic Review and Meta-analysis. Am J Rhinol Allergy. 2021 Mar 1;35(2):195–205. doi:10.1177/1945892420946254

2. Croy I, Nordin S, Hummel T. Olfactory Disorders and Quality of Life—An Updated Review. Chem Senses. 2014 Mar 1;39(3):185–94. doi:10.1093/chemse/bjt072

3. Aschenbrenner K, Hummel C, Teszmer K, Krone F, Ishimaru T, Seo HS, et al. The Influence of Olfactory Loss on Dietary Behaviors. The Laryngoscope. 2008;118(1):135–44. doi:10.1097/MLG.0b013e318155a4b9

4. Rawal S, Duffy VB, Berube L, Hayes JE, Kant AK, Li CM, et al. Self-Reported Olfactory Dysfunction and Diet Quality: Findings from the 2011–2014 National Health and Nutrition Examination Survey (NHANES). Nutrients. 2021 Dec;13(12):12. doi:10.3390/nu13124561

5. Stankevice D, Fjaeldstad A, Ovesen T. Smell and taste disorders in childhood: Diagnostic challenges and significant impacts on a child’s well-being. Int J Pediatr Otorhinolaryngol. 2024 Sep.

6. Lee L, Luke L, Boak D, Philpott C. Impact of olfactory disorders on personal safety and well-being: a cross-sectional observational study. Eur Arch Otorhinolaryngol. 2024;281(7):3639–47. doi:10.1007/s00405-024-08529-9 PubMed PMID: 38396298; PubMed Central PMCID: PMC11211102.

7. Hauser LJ, Jensen EL, Mirsky DM, Chan KH. Pediatric anosmia: A case series. Int J Pediatr Otorhinolaryngol. 2018 Jul;110:135–9. doi:10.1016/j.ijporl.2018.05.011 PubMed PMID: 29859575.

8. Schriever VA, Hummel T. Etiologies of olfactory dysfunction in a pediatric population: based on a retrospective analysis of data from an outpatient clinic. Eur Arch Otorhinolaryngol. 2020;277(11):3213–6. doi:10.1007/s00405-020-06087-4 PubMed PMID: 32488374; PubMed Central PMCID: PMC8249255.

9. Payandeh JE, Motamed M, Kirubalingam K, Chadha NK. Olfactory Dysfunction in Children: A Scoping Review. Otolaryngol Neck Surg. 2023;169(6):1399–408. doi:10.1002/ohn.415

10. Dalton P, Mennella JA, Cowart BJ, Maute C, Pribitkin EA, Reilly JS. Evaluating the Prevalence of Olfactory Dysfunction in a Pediatric Population. Ann N Y Acad Sci. 2009 Jul;1170:537–42. doi:10.1111/j.1749-6632.2009.03919.x PubMed PMID: 19686190; PubMed Central PMCID: PMC3046421.

11. Cameron EL. Olfactory perception in children. World J Otorhinolaryngol - Head Neck Surg. 2018 Mar;4(1):57–66. doi:10.1016/j.wjorl.2018.02.002 PubMed PMID: 30035263; PubMed Central PMCID: PMC6051253.

12. Whitcroft KL, Alobid I, Altundag A, Andrews P, Carrie S, Fahmy M, et al. International clinical assessment of smell: An international, cross-sectional survey of current practice in the assessment of olfaction. Clin Otolaryngol. 2024;49(2):220–34. doi:10.1111/coa.14123

13. Harris PA, Taylor R, Minor BL, Elliott V, Fernandez M, O’Neal L, et al. The REDCap consortium: Building an international community of software platform partners. J Biomed Inform. 2019 Jul;95:103208. doi:10.1016/j.jbi.2019.103208 PubMed PMID: 31078660; PubMed Central PMCID: PMC7254481.

14. Harris PA, Taylor R, Thielke R, Payne J, Gonzalez N, Conde JG. Research electronic data capture (REDCap)--a metadata-driven methodology and workflow process for providing translational research informatics support. J Biomed Inform. 2009 Apr;42(2):377–81. doi:10.1016/j.jbi.2008.08.010 PubMed PMID: 18929686; PubMed Central PMCID: PMC2700030.

15. Petrucci B, Okerosi S, Patterson RH, Hobday SB, Salano V, Waterworth CJ, et al. The Global Otolaryngology–Head and Neck Surgery Workforce. JAMA Otolaryngol Neck Surg. 2023 Oct 1;149(10):904–11. doi:10.1001/jamaoto.2023.2339

16. Nuss S, Nakku D, Pandey A, Elwell Z, Fei C, Zhang D, Srinivasan T, et al. Otolaryngology–Head and Neck Surgery Training and Service Delivery: An International Survey. Laryngoscope Investig Otolaryngol. 2025 Feb 14;10(1):e70096. doi:10.1002/lio2.70096 PubMed PMID: 39958946; PubMed Central PMCID: PMC11826220.

17. Shu CH, Lee PO, Lan MY, Lee YL. Factors affecting the impact of olfactory loss on the quality of life and emotional coping ability. Rhinology. 2011 Aug;49(3):337–41. doi:10.4193/Rhino10.130 PubMed PMID: 21858266.

18. Croy I, Nordin S, Hummel T. Olfactory Disorders and Quality of Life—An Updated Review. Chem Senses. 2014 Mar 1;39(3):185–94. doi:10.1093/chemse/bjt072

19. Croy I, Negoias S, Novakova L, Landis BN, Hummel T. Learning about the Functions of the Olfactory System from People without a Sense of Smell. PLoS ONE. 2012 Mar 21;7(3):e33365. doi:10.1371/journal.pone.0033365 PubMed PMID: 22457756; PubMed Central PMCID: PMC3310072.

20. Croy I, Bojanowski V, Hummel T. Men without a sense of smell exhibit a strongly reduced number of sexual relationships, women exhibit reduced partnership security – a reanalysis of previously published data. Biol Psychol. 2013 Feb;92(2):292–4. doi:10.1016/j.biopsycho.2012.11.008 PubMed PMID: 23178326.

21. Hugh SC, Siu J, Hummel T, Forte V, Campisi P, Papsin BC, et al. Olfactory testing in children using objective tools: comparison of Sniffin’ Sticks and University of Pennsylvania Smell Identification Test (UPSIT). J Otolaryngol - Head Neck Surg J Oto-Rhino-Laryngol Chir Cervico-Faciale. 2015 Mar 1;44(1):10. doi:10.1186/s40463-015-0061-y PubMed PMID: 25890082; PubMed Central PMCID: PMC4359791.

22. Cameron EL, Doty RL. Odor identification testing in children and young adults using the smell wheel. Int J Pediatr Otorhinolaryngol. 2013 Mar;77(3):346–50. doi:10.1016/j.ijporl.2012.11.022 PubMed PMID: 23246420.

23. Gellrich J, Lohrer EC, Hummel T, Schriever VA. Olfactory Dysfunction in Children and Adolescents—A Diagnostic Pathway. Neuropediatrics. 2025 Jan 21;56:215–20. doi:10.1055/a-2509-8547

24. Mariño-Sánchez F, Valls-Mateus M, Fragola C, de Los Santos G, Aguirre A, Alonso J, et al. Pediatric Barcelona Olfactory Test C 6 (pBOT-6): Validation of a Combined Odor Identification and Threshold Screening Test in Healthy Spanish Children and Adolescents. J Investig Allergol Clin Immunol. 2020;30(6):439–47. doi:10.18176/jiaci.0451 PubMed PMID: 31530512.

25. Schriever VA, Agosin E, Altundag A, Avni H, Cao Van H, Cornejo C, et al. Development of an International Odor Identification Test for Children: The Universal Sniff Test. J Pediatr. 2018 Jul 1;198:265–272.e3. doi:10.1016/j.jpeds.2018.03.011

26. Dalton P, Mennella JA, Maute C, Castor SM, Silva-Garcia A, Slotkin J, et al. Development of a Test to Evaluate Olfactory Function in a Pediatric Population. The Laryngoscope. 2011 Sep;121(9):1843–50. doi:10.1002/lary.21928 PubMed PMID: 22024835; PubMed Central PMCID: PMC3800736.

27. Mahmut MK, Pieniak M, Resler K, Schriever VA, Haehner A, Oleszkiewicz A. Olfactory training in 8-year-olds increases odour identification ability: a preliminary study. Eur J Pediatr. 2021 Jul;180(7):2049–53. doi:10.1007/s00431-021-03970-y PubMed PMID: 33566158.

28. Pieniak M, Seidel K, Oleszkiewicz A, Gellrich J, Karpinski C, Fitze G, et al. Olfactory training effects in children after mild traumatic brain injury. Brain Inj. 2023 Sep 19;37(11):1272–84. doi:10.1080/02699052.2023.2237889 PubMed PMID: 37486172.

29. Stankevice D, Fjaeldstad AW, Ovesen T. Smell and taste disorders in childhood: Diagnostic challenges and significant impacts on a child’s well-being. Int J Pediatr Otorhinolaryngol. 2024 Sep 1;184:112081. doi:10.1016/j.ijporl.2024.112081

30. Stull MR, Harshman SG, Kuhnle M, Wons O, Misra M, Eddy K, et al. Olfactory Performance in Youth With Full and Subthreshold Avoidant/Restrictive Food Intake Disorder. J Endocr Soc. 2021 May 3;5(Suppl 1):A630–1. doi:10.1210/jendso/bvab048.1285 PubMed PMID: null; PubMed Central PMCID: PMC8090413.

31. Demattè ML, Endrizzi I, Gasperi F. Food neophobia and its relation with olfaction. Front Psychol. 2014 Feb 17;5:127. doi:10.3389/fpsyg.2014.00127 PubMed PMID: 24596565; PubMed Central PMCID: PMC3925843.

32. Sorokowska A, Chabin D, Kamieńska A, Barszcz S, Byczyńska K, Fuławka K, et al. Olfactory performance and odor liking are negatively associated with food neophobia in children aged between 3 and 9 years. Nutr J. 2024 Sep 11;23(1):105. doi:10.1186/s12937-024-01011-6

33. Hummel T, T. Liu D, A. Müller C, A. Stuck B, Welge-Lüssen A, Hähner A. Olfactory Dysfunction: Etiology, Diagnosis, and Treatment. Dtsch Ärztebl Int. 2023 Mar;120(9):146–54. doi:10.3238/arztebl.m2022.0411 PubMed PMID: 36647581; PubMed Central PMCID: PMC10198165.

34. Sattar Othman M, Hu K, Davidson J, Kirubalingam K, Graham ME, Coyle P, et al. The Utilization of Artificial Intelligence by Pediatric Otolaryngology Surgeons in Professional Practice. J Otolaryngol - Head Neck Surg J Oto-Rhino-Laryngol Chir Cervico-Faciale. 2026;55:19160216251411838. doi:10.1177/19160216251411838 PubMed PMID: 41504171; PubMed Central PMCID: PMC12783579.

35. Patel A, Dzioba A, Hong P, Husein M, Strychowsky J, You P, et al. Changes to the practice of pediatric otolaryngology as a consequence of the COVID-19 pandemic. Int J Pediatr Otorhinolaryngol. 2022 Feb 1;153:111021. doi:10.1016/j.ijporl.2021.111021

36. Spencer GM, Wilson CA, Davidson J, Strychowsky JE, Lawlor CM, Burns H, et al. Global tonsillectomy practice patterns – A survey study of pediatric otolaryngologists. Int J Pediatr Otorhinolaryngol. 2025 Apr 1;191:112276. doi:10.1016/j.ijporl.2025.112276

37. Spencer GM, Wilson CA, Davidson J, Strychowsky JE, Lawlor CM, Burns H, et al. International Practice Variation in Post-Tonsillectomy Hemorrhage: A Survey Study of Pediatric Otolaryngologists. World J Otorhinolaryngol - Head Neck Surg. n/a(n/a). doi:10.1002/wjo2.70068

38. Manouchehri K, Zahabi S, Davidson J, Wilson CA, Lawlor C, Graham ME. Knowledge and attitudes surrounding breastfeeding in pediatric otolaryngology: A survey study. Int J Pediatr Otorhinolaryngol. 2024 Jan 1;176:111774. doi:10.1016/j.ijporl.2023.111774

39. Maniaci A, Chiesa Estomba C, Fakhry N, Vaira LA, Remacle M, Cammaroto G, et al. Influence of Otolaryngological Subspecialties on Perception of Transoral Robotic Surgery: An International YO-IFOS Survey. J Pers Med. 2023 Dec;13(12):1717. doi:10.3390/jpm13121717

