## Supplementary Tables for "Global practices in paediatric olfactory dysfunction: a cross-sectional survey of paediatric ENT surgeons"

| **Primary tools for assessment of olfactory function for ages 0-2 years. N (%)** | | | | | | | | | | | |
| --- | --- | --- | --- | --- | --- | --- | --- | --- | --- | --- | --- |
| **Categories** | **Africa** | **Asia** | **Australia** | **Europe** | **Middle East** | **North America** | **South America** | **UK** | **USA** | **Total** | **Chi-square p-value** |
| Behavioural cues | 0 (0) | 0 (0) | 0 (0) | 0 (0) | 0 (0) | 0 (0) | 0 (0) | 1 (2.6) | 0 (0) | 1 (0.6) | 0.914 |
| Parental report | 0 (0) | 0 (0) | 0 (0) | 0 (0) | 0 (0) | 0 (0) | 0 (0) | 1 (2.6) | 0 (0) | 1 (0.6) | 0.914 |
| Other | 0 (0) | 0 (0) | 0 (0) | 0 (0) | 0 (0) | 0 (0) | 0 (0) | 0 (0) | 0 (0) | 0 (0) | - |
| Not assessed | 0 (0) | 0 (0) | 0 (0) | 0 (0) | 0 (0) | 0 (0) | 0 (0) | 0 (0) | 0 (0) | 0 (0) | - |
| **Primary tools for assessment of olfactory function for ages 3-6 years. N (%)** | | | | | | | | | | | |
| **Categories** | **Africa** | **Asia** | **Australia** | **Europe** | **Middle East** | **North America** | **South America** | **UK** | **USA** | **Total** | **Chi-square p-value** |
| U-Sniff | 0 (0) | 0 (0) | 0 (0) | 0 (0) | 0 (0) | 0 (0) | 0 (0) | 2 (5.1) | 0 (0) | 2 (1.2) | 0.576 |
| Modified Sniffin’ kids | 0 (0) | 0 (0) | 1 (3.7) | 2 (8.7) | 0 (0) | 0 (0) | 0 (0) | 0 (0) | 0 (0) | 3 (1.8) | 0.351 |
| Odour identification games | 0 (0) | 1 (6.3) | 0 (0) | 3 (13.0) | 0 (0) | 0 (0) | 0 (0) | 0 (0) | 0 (0) | 4 (2.4) | 0.053 |
| UPSIT/BSIT without paediatric norms | 0 (0) | 1 (6.3) | 0 (0) | 0 (0) | 0 (0) | 0 (0) | 0 (0) | 1 (2.6) | 0 (0) | 2 (1.2) | 0.711 |
| Not assessed | 0 (0) | 0 (0) | 1 (3.7) | 0 (0) | 0 (0) | 0 (0) | 0 (0) | 0 (0) | 0 (0) | 1 (0.6) | 0.734 |
| **Primary tools for assessment of olfactory function for ages 7-12 years. N (%)** | | | | | | | | | | | |
| **Categories** | **Africa** | **Asia** | **Australia** | **Europe** | **Middle East** | **North America** | **South America** | **UK** | **USA** | **Total** | **Chi-square p-value** |
| U-Sniff | 0 (0) | 0 (0) | 1 (3.7) | 2 (8.7) | 0 (0) | 0 (0) | 1 (14.3) | 2 (5.1) | 1 (4.5) | 7 (4.2) | 0.737 |
| Sniffin’ kids | 0 (0) | 0 (0) | 1 (3.7) | 3 (13) | 0 (0) | 0 (0) | 1 (14.3) | 2 (5.1) | 1 (4.5) | 8 (4.8) | 0.498 |
| Standard Sniffin’ Sticks | 0 (0) | 1 (6.3) | 0 (0) | 3 (13) | 0 (0) | 0 (0) | 1 (14.3) | 0 (0) | 1 (4.5) | 6 (3.6) | 0.138 |
| Paediatric Smell Wheel | 0 (0) | 0 (0) | 0 (0) | 0 (0) | 0 (0) | 0 (0) | 0 (0) | 1 (2.6) | 4 (18.2) | 5 (3.0) | **0.008** |
| UPSIT/BSIT without paediatric norms | 0 (0) | 2 (12.5) | 1 (3.7) | 0 (0) | 1 (5.6) | 0 (0) | 0 (0) | 3 (7.7) | 7 (31.8) | 14 (8.4) | **0.007** |
| Not assessed | 0 (0) | 1 (6.3) | 1 (3.7) | 0 (0) | 0 (0) | 0 (0) | 0 (0) | 0 (0) | 0 (0) | 2 (1.2) | 0.604 |
| **Primary tools for assessment of olfactory function for over 12 years. N (%)** | | | | | | | | | | | |
| **Categories** | **Africa** | **Asia** | **Australia** | **Europe** | **Middle East** | **North America** | **South America** | **UK** | **USA** | **Total** | **Chi-square p-value** |
| Adult protocols with adaptations | 0 (0) | 5 (31.3) | 5 (18.5) | 3 (13) | 3 (16.7) | 1 (10) | 1 (14.3) | 4 (10.3) | 11 (50) | 33 (19.8) | **0.015** |
| Hormonal assessment | 0 (0) | 2 (12.5) | 2 (7.4) | 0 (0) | 0 (0) | 1 (10) | 0 (0) | 2 (5.1) | 0 (0) | 7 (4.2) | 0.468 |
| Not assessed | 0 (0) | 2 (12.5) | 0 (0) | 0 (0) | 1 (5.6) | 0 (0) | 0 (0) | 2 (5.1) | 0 (0) | 5 (3.0) | 0.351 |

Supplementary Table 1. Primary tools used by clinicians for assessment of olfactory function in children across different age groups (0–2, 3–6, 7–12, and over 12 years), by geographic region. Tools include psychophysical tests (e.g., U-Sniff, Sniffin’ Sticks), informal assessments (e.g., behavioural cues, parental report), and adapted adult protocols. Not all respondents assess olfaction at all age ranges. Chi-square p-values represent statistical comparisons of usage patterns by region.

| **Reported Barriers to Objective Olfactory Testing in Clinical Practice. N (%)** | | | | | | | | | | | |
| --- | --- | --- | --- | --- | --- | --- | --- | --- | --- | --- | --- |
| **Categories** | **Africa** | **Asia** | **Australia** | **Europe** | **Middle East** | **North America** | **South America** | **UK** | **USA** | **Total** | **Chi-square p-value** |
| N/A – I use smell tests routinely | 0 (0) | 0 (0) | 1 (3.7) | 4 (17.4) | 0 (0) | 1 (10) | 1 (14.3) | 4 (10.3) | 6 (27.3) | 17 (10.2) | 0.079 |
| Insufficient time | 0 (0) | 5 (31.3) | 7 (25.9) | 7(30.4) | 4 (22.2) | 3 (30) | 3 (42.9) | 12 (30.8) | 9 (40.9) | 50 (29.9) | 0.789 |
| Insufficient staff | 0 (0) | 4 (25) | 3 (11.1) | 6 (26.1) | 1 (5.6) | 0 (0) | 0 (0) | 9 (23.1) | 3 (13.6) | 26 (15.6) | 0.226 |
| Insufficient experience/training | 3 (60) | 8 (50) | 15 (55.6) | 7 (30.4) | 13 (72.2) | 4 (40) | 3 (42.9) | 13 (33.3) | 8 (36.4) | 74 (44.3) | 0.146 |
| Funding limitations (from your hospital/institution) | 0 (0) | 2 (12.5) | 11 (40.7) | 6 (26.1) | 6 (33.3) | 3 (30) | 5 (71.4) | 11 (28.2) | 3 (13.6) | 47 (28.1) | 0.057 |
| Funding limitations (from patient/insurance) | 1 (20) | 4 (25) | 3 (11.1) | 1 (4.3) | 2 (11.1) | 1 (10) | 3 (42.9) | 2 (5.1) | 1 (4.5) | 18 (10.8) | 0.07 |
| Lack of reliable power supply | 0 (0) | 2 (12.5) | 0 (0) | 0 (0) | 3 (16.7) | 0 (0) | 0 (0) | 1 (2.6) | 0 (0) | 6 (3.6) | **0.04** |
| Poor access to equipment maintenance | 0 (0) | 2 (12.5) | 6 (22.2) | 6 (26.1) | 5 (27.8) | 0 (0) | 2 (28.6) | 5 (12.8) | 0 (0) | 26 (15.6) | 0.114 |
| Lack of validation data for the population you treat | 0 (0) | 6 (37.5) | 5 (18.5) | 5 (21.7) | 3 (16.7) | 1 (10) | 1 (14.3) | 8 (20.5) | 6 (27.3) | 35 (21) | 0.678 |
| Cultural mismatch of odourants | 0 (0) | 3 (18.8) | 0 (0) | 2 (8.7) | 3 (16.7) | 0 (0) | 2 (28.6) | 2 (5.1) | 0 (0) | 12 (7.2) | **0.047** |
| History/Examination is sufficient | 0 (0) | 2 (12.5) | 1 (3.7) | 1 (4.3) | 0 (0) | 0 (0) | 0 (0) | 3 (7.7) | 0 (0) | 7 (4.2) | 0.573 |
| Will not impact management | 0 (0) | 0 (0) | 6 (22.2) | 3 (13) | 4 (22.2) | 3 (30) | 0 (0) | 6 (15.4) | 2 (9.1) | 24 (14.4) | 0.302 |
| **Reported Barriers to Early Diagnosis of Paediatric Olfactory Dysfunction by Region. N (%)** | | | | | | | | | | | |
| **Categories** | **Africa** | **Asia** | **Australia** | **Europe** | **Middle East** | **North America** | **South America** | **UK** | **USA** | **Total** | **Chi-square p-value** |
| Lack of caregiver awareness | 12 (40) | 12 (75) | 13 (48.1) | 14 (60.9) | 10 (55.6) | 8 (80) | 5 (71.4) | 15 (38.5) | 13 (59.1) | 92 (55.1) | 0.169 |
| Lack of referring clinician/primary care awareness | 3 (60) | 9 (56.3) | 16 (59.3) | 17 (73.9) | 11 (61.1) | 7 (70) | 3 (42.9) | 23 (59) | 11 (50) | 100 (59.9) | 0.842 |
| Normalisation of symptoms | 2 (40) | 6 (37.5) | 8 (29.6) | 7 (30.4) | 10 (55.6) | 5 (50) | 6 (85.7) | 17 (43.7) | 9 (40.9) | 70 (41.9) | 0.247 |
| Insensitive screening tools | 0 (0) | 2 (12.5) | 7 (25.9) | 4 (17.4) | 5 (27.8) | 3 (30) | 1 (14.3) | 6 (15.4) | 3 (13.6) | 31 (18.6) | 0.748 |
| Funding challenges – Hospital/Institution | 0 (0) | 1 (6.3) | 4 (14.8) | 2 (8.7) | 0 (0) | 3 (30) | 3 (42.9) | 9 (23.1) | 1 (4.5) | 23 (13.8) | **0.035** |
| Funding challenges – Patient/Insurance Reimbursement | 0 (0) | 1 (6.3) | 1 (3.7) | 1 (4.3) | 0 (0) | 0 (0) | 3 (42.9) | 1 (2.6) | 1 (4.5) | 8 (4.8) | **0.002** |

Supplementary Table 2. Respondents’ perceptions of the most common barriers to performing objective smell testing in children, as reported across global regions, as well as perceived challenges to timely identification of olfactory dysfunction in children.

| **Frequency of Smell Screening Offered in Pediatric Patients with Select Conditions. N (%)** | | | | | | | | | | | | |
| --- | --- | --- | --- | --- | --- | --- | --- | --- | --- | --- | --- | --- |
| **Condition** | **Categories** | **Africa** | **Asia** | **Australia** | **Europe** | **Middle East** | **North America** | **South America** | **UK** | **USA** | **Total** | **Chi-square p-value** |
| Chronic rhinosinusitis | Routinely | 1 (33.3) | 1 (7.7) | 3 (13.6) | 0 (0) | 0 (0) | 1 (12.5) | 1 (16.7) | 3 (9.1) | 1 (4.8) | 11 (7.7) | 0.129 |
|  | Sometimes | 0 (0) | 3 (23.1) | 2 (9.1) | 2 (10) | 1 (5.9) | 2 (25) | 2 (33.3) | 1 (3) | 2 (9.5) | 15 (10.5) |  |
|  | Only if complaints of OD | 1 (33.3) | 4 (30.8) | 4 (18.2) | 8 (40) | 4 (23.5) | 1 (12.5) | 1 (16.7) | 10 (30.3) | 13 (61.9) | 46 (32.2) |  |
|  | Never | 1 (33.3) | 5 (38.5) | 13 (59.1) | 10 (50) | 12 (70.6) | 4 (50) | 2 (33.3) | 19 (57.6) | 5 (23.8) | 71 (49.7) |  |
| Obstructive nasal pathology (deviated septum/adenoid hypertrophy) | Routinely | 1 (33.3) | 0 (0) | 4 (18.2) | 0 (0) | 1 (5.9) | 0 (0) | 0 (0) | 0 (0) | 0 (0) | 6 (4.3) | **0.001** |
|  | Sometimes | 0 (0) | 3 (23.1) | 0 (0) | 0 (0) | 0 (0) | 0 (0) | 0 (0) | 3 (9.7) | 3 (14.3) | 9 (6.4) |  |
|  | Only if complaints of OD | 1 (33.3) | 5 (38.5) | 4 (18.2) | 10 (52.6) | 2 (11.8) | 2 (25) | 4 (66.7) | 6 (19.4) | 9 (42.9) | 43 (30.7) |  |
|  | Never | 1 (33.3) | 5 (38.5) | 14 (63.6) | 9 (47.4) | 14 (82.4) | 6 (75) | 2 (33.3) | 22 (71) | 9 (42.9) | 82 (58.6) |  |
| Allergic Rhinitis | Routinely | 0 (0) | 1 (7.7) | 4 (18.2) | 0 (0) | 1 (5.9) | 0 (0) | 0 (0) | 1 (3.2) | 1 (4.8) | 8 (5.7) | **0.05** |
|  | Sometimes | 1 (33.3) | 2 (15.4) | 1 (4.5) | 2 (10.5) | 0 (0) | 0 (0) | 0 (0) | 2 (6.5) | 2 (9.5) | 10 (7.1) |  |
|  | Only if complaints of OD | 1 (33.3) | 6 (46.2) | 3 (13.6) | 8 (42.1) | 2 (11.8) | 2 (22.2) | 4 (66.7) | 7 (22.6) | 11 (52.4) | 44 (31.2) |  |
|  | Never | 1 (33.3) | 4 (30.8) | 14 (63.6) | 9 (47.4) | 14 (82.4) | 7 (77.8) | 2 (33.3) | 21 (67.7) | 7 (33.3) | 79 (56) |  |
| Post-viral perceived OD | Routinely | 1 (33.3) | 2 (14.3) | 4 (18.2) | 7 (36.8) | 3 (17.6) | 1 (11.1) | 2 (33.3) | 7 (21.9) | 9 (42.9) | 36 (25.2) | 0.092 |
|  | Sometimes | 0 (0) | 3 (21.4) | 4 (18.2) | 2 (10.5) | 1 (5.9) | 3 (33.3) | 2 (33.3) | 2 (6.3) | 9 (42.9) | 26 (18.2) |  |
|  | Only if complaints of OD | 1 (33.3) | 5 (35.7) | 7 (31.8) | 6 (31.6) | 6 (35.3) | 1 (11.1) | 1 (16.7) | 9 (28.1) | 1 (4.8) | 37 (25.9) |  |
|  | Never | 1 (33.3) | 4 (28.6) | 7 (31.8) | 4 (21.1) | 7 (41.2) | 4 (44.4) | 1 (16.7) | 14 (43.8) | 2 (9.5) | 44 (30.8) |  |
| Primary ciliary dyskinesia | Routinely | 1 (33.3) | 4 (28.6) | 2 (9.1) | 0 (0) | 1 (5.9) | 1 (11.1) | 2 (33.3) | 2 (6.5) | 0 (0) | 13 (9.2 | **0.039** |
|  | Sometimes | 0 (0) | 3 (21.4) | 3 (13.6) | 4 (21.1) | 1 (5.9) | 1 (11.1) | 0 (0) | 5 (16.1) | 4 (19) | 21 (14.8) |  |
|  | Only if complaints of OD | 1 (33.3) | 3 (21.4) | 6 (27.3) | 8 (42.1) | 5 (29.4) | 1 (11.1) | 2 (33.3) | 4 (12.9) | 12 (57.1) | 42 (29.6) |  |
|  | Never | 1 (33.3) | 4 (28.6) | 11 (50) | 7 (36.8) | 10 (58.8) | 6 (66.7) | 2 (33.3) | 20 (64.5) | 5 (23.8) | 66 (46.5) |  |
| Cystic fibrosis | Routinely | 1 (33.3) | 3 (23.1) | 2 (9.1) | 0 (0) | 1 (5.9) | 1 (11.1) | 2 (33.3) | 1 (3.2) | 0 (0) | 11 (7.9) | **0.037** |
|  | Sometimes | 0 (0) | 3 (23.1) | 4 (18.2) | 3 (16.7) | 1 (5.9) | 2 (22.2) | 0 (0) | 6 (19.4) | 3 (14.3) | 22 (15.7) |  |
|  | Only if complaints of OD | 1 (33.3) | 3 (23.1) | 5 (22.7) | 7 (38.9) | 5 (29.4) | 1 (11.1) | 2 (33.3) | 4 (12.9) | 13 (61.9) | 41 (29.3) |  |
|  | Never | 1 (33.3) | 4 (30.8) | 11 (50) | 8 (44.4) | 10 (58.8) | 5 (55.6) | 2 (33.3) | 20 (64.5) | 5 (23.8) | 66 (47.1) |  |
| Traumatic brain injury | Routinely | 1 (33.3) | 2 (14.3) | 3 (13.6) | 6 (31.6) | 1 (5.9) | 0 (0) | 2 (33.3) | 5 (15.6) | 1 (4.8) | 21 (14.7) | 0.396 |
|  | Sometimes | 0 (0) | 2 (14.3) | 3 (13.6) | 1 (5.3) | 2 (11.8) | 2 (22.2) | 1 (16.7) | 1 (3.1) | 3 (14.3) | 15 (10.5) |  |
|  | Only if complaints of OD | 1 (33.3) | 5 (35.7) | 9 (40.9) | 4 (21.1) | 6 (35.3) | 2 (22.2) | 2 (33.3) | 8 (25) | 12 (57.1) | 49 (34.3) |  |
|  | Never | 1 (33.3) | 5 (35.7) | 7 (31.8) | 8 (42.1) | 8 (47.1) | 5 (55.6) | 1 (16.7) | 18 (56.3) | 5 (23.8) | 58 (40.6) |  |
| 22q11 deletion syndrome | Routinely | 0 (0) | 0 (0) | 2 (9.1) | 0 (0) | 0 (0) | 0 (0) | 0 (0) | 0 (0) | 0 (0) | 2 (1.4) | 0.226 |
|  | Sometimes | 1 (33.3) | 3 (25) | 3 (13.6) | 1 (5.6) | 1 (5.9) | 0 (0) | 1 (16.7) | 3 (9.7) | 2 (9.5) | 15 (10.9) |  |
|  | Only if complaints of OD | 1 (33.3) | 5 (41.7) | 6 (27.3) | 9 (50) | 6 (35.3) | 1 (12.5) | 3 (50) | 8 (25.8) | 12 (57.1) | 51 (37) |  |
|  | Never | 1 (33.3) | 4 (33.3) | 11 (50) | 8 (44.4) | 10 (58.8) | 7 (87.5) | 2 (33.3) | 20 (64.5) | 7 (33.3) | 70 (50.7) |  |
| Trisomy 21 | Routinely | 0 (0) | 0 (0) | 2 (9.1) | 0 (0) | 1 (5.9) | 0 (0) | 0 (0) | 0 (0) | 0 (0) | 3 (2.2) | 0.158 |
|  | Sometimes | 0 (0) | 2 (15.4) | 1 (4.5) | 0 (0) | 0 (0) | 0 (0) | 1 (16.7) | 2 (6.7) | 1 (5) | 7 (5) |  |
|  | Only if complaints of OD | 2 (66.7) | 6 (46.2) | 7 (31.8) | 9 (47.4) | 4 (23.5) | 0 (0) | 3 (50) | 8 (26.7) | 11 (55) | 50 (36) |  |
|  | Never | 1 (33.3) | 5 (38.5) | 12 (54.5) | 10 (52.6) | 12 (70.6) | 9 (100) | 2 (33.3) | 20 (66.7) | 8 (40) | 79 (56.8) |  |
| CHARGE syndrome | Routinely | 0 (0) | 0 (0) | 2 (9.5) | 3 (16.7) | 1 (5.9) | 0 (0) | 0 (0) | 0 (0) | 0 (0) | 6 (4.4) | **0.043** |
|  | Sometimes | 1 (33.3) | 2 (15.4) | 2 (9.5) | 1 (5.6) | 0 (0) | 0 (0) | 1 (16.7) | 1 (3.2) | 1 (5) | 9 (6.6) |  |
|  | Only if complaints of OD | 1 (33.3) | 6 (46.2) | 6 (28.6) | 6 (33.3) | 5 (29.4) | 0 (0) | 3 (50) | 9 (29) | 13 (65) | 49 (35.8) |  |
|  | Never | 1 (33.3) | 5 (38.5) | 11 (52.4) | 8 (44.4) | 11 (64.7) | 8 (100) | 2 (33.3) | 21 (67.7) | 6 (30) | 73 (53.3) |  |
| Kallman syndrome/hypogonadotropic hypognadism | Routinely | 0 (0) | 6 (54.5) | 3 (15.8) | 8 (44.4) | 5 (31.3) | 4 (44.4) | 2 (33.3) | 5 (16.1) | 7 (36.8) | 40 (30.5) | 0.368 |
|  | Sometimes | 0 (0) | 0 (0) | 0 (0) | 1 (5.6) | 2 (12.5) | 1 (11.1) | 0 (0) | 2 (6.5) | 1 (5.3) | 7 (5.3) |  |
|  | Only if complaints of OD | 1 (50) | 3 (27.3) | 7 (36.8) | 3 (16.7) | 3 (18.8) | 0 (0) | 2 (33.3) | 7 (22.6) | 8 (42.1) | 34 (26) |  |
|  | Never | 1 (50) | 2 (18.2) | 9 (47.4) | 6 (33.3) | 6 (37.5) | 4 (44.4) | 2 (33.3) | 17 (54.8) | 3 (15.8) | 50 (38.2) |  |
| Bardet-Biedl syndrome | Routinely | 0 (0) | 0 (0) | 0 (0) | 2 (11.1) | 1(5.9) | 0 (0) | 0 (0) | 1 (3.2) | 0 (0) | 4 (3) | 0.115 |
|  | Sometimes | 0 (0) | 2 (16.7) | 2 (9.5) | 3 (16.7) | 1 (5.9) | 1 (11.1) | 1 (16.7) | 0 (0) | 4 (22.2) | 14 (10.4) |  |
|  | Only if complaints of OD | 2 (66.7) | 6 (50) | 7 (33.3) | 7 (38.9) | 4 (23.5) | 1 (11.1) | 3 (50) | 7 (22.6) | 10 (55.6) | 47 (34.8) |  |
|  | Never | 1 (33.3) | 4 (33.3) | 12 (57.1) | 6 (33.3) | 11 (64.7) | 7 (77.8) | 2 (33.3) | 23 (74.2) | 4 (22.2) | 70 (51.9) |  |
| Child receiving chemotherapy | Routinely | 0 (0) | 1 (7.1) | 1 (4.8) | 2 (10.5) | 0 (0) | 0 (0) | 2 (33.3) | 0 (0) | 1 (5.6) | 7 (5.1) | 0.071 |
|  | Sometimes | 0 (0) | 1 (7.1) | 2 (9.5) | 1 (5.3) | 2 (11.8) | 0 (0) | 0 (0) | 0 (0) | 1 (5.6) | 7 (5.1) |  |
|  | Only if complaints of OD | 2 (66.7) | 7 (50) | 7 (33.3) | 8 (42.1) | 6 (35.3) | 1 (11.1) | 2 (33.3) | 9 (30) | 12 (66.7) | 54 (39.4) |  |
|  | Never | 1 (33.3) | 5 (35.7) | 11 (52.4) | 8 (42.1) | 9 (52.9) | 8 (88.9) | 2 (33.3) | 21 (70) | 4 (22.2) | 69 (50.4) |  |
| Autism spectrum disorder | Routinely | 0 (0) | 0 (0) | 0 (0) | 1 (5.3) | 0 (0) | 0 (0) | 0 (0) | 0 (0) | 0 (0) | 1 (0.7) | **0.043** |
|  | Sometimes | 0 (0) | 2 (15.4) | 2 (9.5) | 1 (5.3) | 1 (5.9) | 0 (0) | 1 (16.7) | 0 (0) | 0 (0) | 7 (5.1) |  |
|  | Only if complaints of OD | 2 (66.7) | 6 (46.2) | 7 (33.3) | 7 (36.8) | 4 (23.5) | 0 (0) | 2 (33.3) | 5 (16.7) | 12 (66.7) | 45 (33.1) |  |
|  | Never | 1 (33.3) | 5 (38.5) | 12 (57.1) | 10 (52.6) | 12 (70.6) | 9 (100) | 3 (50) | 25 (83.3) | 6 (33.3) | 83 (61) |  |
| Eating disorders | Routinely | 0 (0) | 0 (0) | 0 (0) | 3 (15.8) | 0 (0) | 0 (0) | 1 (16.7) | 0 (0) | 0 (0) | 4 (3) | **0.003** |
|  | Sometimes | 0 (0) | 3 (25) | 1 (4.8) | 1 (5.3) | 1 (6.3) | 0 (0) | 0 (0) | 0 (0) | 3 (16.7) | 9 (6.7) |  |
|  | Only if complaints of OD | 2 (66.7) | 4 (33.3) | 8 (38.1) | 7 (36.8) | 5 (31.3) | 1 (11.1) | 2 (33.3) | 5 (16.7) | 10 (55.6) | 44 (32.8) |  |
|  | Never | 1 (33.3) | 5 (41.7) | 12 (57.1) | 8 (42.1) | 10 (62.5) | 8 (88.9) | 3 (50) | 25 (83.3) | 5 (27.8) | 77 (57.5) |  |
| **Management Strategies Utilized for Pediatric Congenital Anosmia. N (%)** | | | | | | | | | | | | |
|  | **Categories** | **Africa** | **Asia** | **Australia** | **Europe** | **Middle East** | **North America** | **South America** | **UK** | **USA** | **Total** | **Chi-square p-value** |
|  | Full neurological examination | 2 (40) | 10 (62.5) | 13 (48.1) | 12 (52.2) | 11 (61.1) | 4 (40) | 4 (57.1) | 16 (41) | 11 (50) | 83 (49.7) | 0.86 |
|  | Endocrine referral/investigations | 2 (40) | 12 (75) | 12 (44.4) | 14 (60.9) | 8 (44.4) | 4 (40) | 3 (42.9) | 18 (46.2) | 12 (54.5) | 85 (50.9) | 0.564 |
|  | Genetic counselling | 1 (20) | 4 (25) | 2 (7.4) | 10 (43.5) | 7 (38.9) | 2 (20) | 3 (42.9) | 7 (17.9) | 14 (63.6) | 50 (29.9) | **0.001** |
|  | Safety training | 2 (40) | 4 (25) | 10 (37) | 7 (30.4) | 6 (33.3) | 4 (40) | 3 (42.9) | 17 (43.6) | 12 (54.5) | 65 (38.9) | 0.763 |
|  | Puberty delay monitoring | 0 (0) | 1 (6.3) | 2 (7.4) | 6 (26.1) | 2 (11.10 | 1 (10) | 3 (42.9) | 4 (10.3) | 2 (9.1) | 21 (12.6) | 0.145 |
|  | Peer support group advice | 0 (0) | 1 (6.3) | 0 (0) | 2 (8.7) | 1 (5.6) | 0 (0) | 0 (0) | 4 (10.3) | 0 (0) | 8 (4.8) | 0.537 |
|  | No active intervention | 0 (0) | 0 (0) | 3 (11.1) | 3 (13) | 2 (11.1) | 1 (10) | 0 (0) | 2 (5.1) | 1 (4.5) | 12 (7.2) | 0.759 |
|  | N/A – I do not manage children with congenital anosmia in my practice | 2 (40) | 3 (18.8) | 6 (22.2) | 2 (8.7) | 5 (27.8) | 2 (20) | 2 (28.6) | 4 (10.3) | 1 (4.5) | 27 (16.2) | 0.284 |
| **Management Strategies Utilized for Acute Post-Viral Olfactory Dysfunction in Children. N (%)** | | | | | | | | | | | | |
|  | **Categories** | **Africa** | **Asia** | **Australia** | **Europe** | **Middle East** | **North America** | **South America** | **UK** | **USA** | **Total** | **Chi-square p-value** |
|  | Observation | 1 (20) | 8 (50) | 8 (29.6) | 6 (26.1) | 4 (22.2) | 4 (40) | 0 (0) | 16 (41) | 4 (18.2) | 51 (30.5) | 0.2 |
|  | Topical steroids | 3 (60) | 10 (62.5) | 14 (51.9) | 18 (78.3) | 13 (72.2) | 8 (80) | 7 (100) | 28 (71.8) | 14 (63.6) | 115 (68.9) | 0.314 |
|  | Systemic steroids | 1 (20) | 3 (18.8) | 4 (14.8) | 6 (26.1) | 1 (5.6) | 0 (0) | 1 (14.3) | 3 (7.7) | 1 (4.5) | 20 (12) | 0.303 |
|  | Olfactory training | 2 (40) | 5 (31.3) | 9 (33.3) | 14 (60.9) | 4 (22.2) | 3 (30) | 6 (85.7) | 18 (46.2) | 15 (68.2) | 76 (45.5) | **0.014** |
|  | Oral supplements (Omega-3) | 0 (0) | 3 (18.8) | 7 (25.9) | 2 (8.7) | 0 (0) | 1 (10) | 2 (28.6) | 7 (17.9) | 2 (9.1) | 24 (14.4) | 0.276 |
|  | N/A – I do not manage children with post – viral OD in my practice | 1 (20) | 1 (6.3) | 5 (18.5) | 2 (8.7) | 2 (11.1) | 1 (10) | 0 (0) | 0 (0) | 2 (9.1) | 14 (8.4) | 0.341 |

Supplementary Table 3. summarizes clinician-reported practices regarding including (1) the frequency with which smell screening is offered in specific conditions, (2) preferred management strategies for congenital anosmia and (3) preferred management strategies for acute post-viral olfactory dysfunction.

| **Frequency of Multidisciplinary Collaboration in the Management of Non-Syndromic Pediatric Olfactory Dysfunction** | | | | | | | | | | | | |
| --- | --- | --- | --- | --- | --- | --- | --- | --- | --- | --- | --- | --- |
| **Categories** | **Category** | **Africa** | **Asia** | **Australia** | **Europe** | **Middle East** | **North America** | **South America** | **UK** | **USA** | **Total** | **Chi-square p-value** |
| Paediatric neurology | Routinely | 0 (0) | 4 (40) | 1 (4.8) | 2 (11.1) | 3 (20) | 0 (0) | 0 (0) | 1 (3.1) | 1 (5.6) | 12 (9.4) | **0.002** |
|  | Often | 0 (0) | 4 (40) | 4 (19) | 7 (38.9) | 1 (6.7) | 1 (16.7) | 0 (0) | 1 (3.1) | 5 (27.8) | 23 (18.1) |  |
|  | Sometimes | 1(33.3) | 0 (0) | 6 (28.6) | 5 (27.8) | 5 (33.3) | 0 (0) | 1 (25) | 12 (37.5) | 6 (33.3) | 36 (28.3) |  |
|  | Rarely | 0 (0) | 1 (10) | 6 (28.6) | 0 (0) | 5 (33.3) | 1 (16.7) | 3 (75) | 10 (31.3) | 3 (16.7) | 29 (22.8) |  |
|  | Never | 2 (66.7) | 1 (10) | 4 (19) | 4 (22.2) | 1 (6.7) | 4 (66.7) | 0 (0) | 8 (25) | 3 (16.7) | 27 (21.3) |  |
| Paediatric endocrinology | Routinely | 0 (0) | 4 (36.4) | 3 (14.3) | 2 (11.8) | 2 (13.3) | 1 (16.7) | 1 (25) | 2 (6.5) | 2 (10) | 17 (13.3) | 0.516 |
|  | Often | 0 (0) | 3 (27.3) | 1 (4.8) | 2 (11.8) | 1 (6.7) | 0 (0) | 0 (0) | 3 (9.7) | 4 (20) | 14 (10.9) |  |
|  | Sometimes | 1 (33.3) | 3 (27.3) | 10 (47.6) | 8 (47.1) | 5 (33.3) | 2 (33.3) | 0 (0) | 12 (38.7) | 8 (40) | 49 (38.3) |  |
|  | Rarely | 0 (0) | 0 (0) | 4 (19) | 2 (11.8) | 4 (26.7) | 0 (0) | 2 (50) | 7 (22.6) | 3 (15) | 22 (17.2) |  |
|  | Never | 2 (6.7) | 1 (9.1) | 3 (14.3) | 3 (17.6) | 3 (20) | 3 (50) | 1 (25) | 7 (22.6) | 3 (15) | 26 (20.3) |  |
| Neuropsychology | Routinely | 0 (0) | 1 (12.5) | 1 (4.8) | 0 (0) | 1 (7.7) | 0 (0) | 0 (0) | 0 (0) | 0 (0) | 3 (2.5) | 0.255 |
|  | Often | 0 (0) | 1 (12.5) | 0 (0) | 3 (17.6) | 0 (0) | 0 (0) | 0 (0) | 0 (0) | 2 (11.1) | 6 (5) |  |
|  | Sometimes | 0 (0) | 2 (25) | 5 (23.8) | 3 (17.6) | 5 (38.5) | 0 (0) | 0 (0) | 4 (13.8) | 6 (33.3) | 25 (21) |  |
|  | Rarely | 1 (33.3) | 1 (12.5) | 3 (14.3) | 4 (23.5) | 4 (30.8) | 1 (16.7) | 2 (50) | 5 (17.2) | 5 (27.8) | 26 (21.8) |  |
|  | Never | 2 (66.7) | 3 (37.5) | 12 (57.1) | 7 (41.2) | 3 (23.1) | 5 (83.3) | 2 (50) | 20 (69) | 5 (27.8) | 59 (49.6) |  |
| Clinical genetics | Routinely | 0 (0) | 3 (33.3) | 1 (4.8) | 1 (5.6) | 2 (13.3) | 1 (16.7) | 0 (0) | 0 (0) | 3 (15.8) | 11 (8.8) | 0.082 |
|  | Often | 0 (0) | 3 (33.3) | 1 (4.8) | 2 (11.1) | 2 (13.3) | 0 (0) | 1 (25) | 1 (3.3) | 2 (10.5) | 12 (9.6) |  |
|  | Sometimes | 0 (0) | 2 (22.2) | 5 (23.8) | 7 (38.9) | 4 (26.7) | 0 (0) | 0 (0) | 8 (26.7) | 9 (47.4) | 35 (28) |  |
|  | Rarely | 1 (33.3) | 0 (0) | 8 (38.1) | 3 (16.7) | 3 (20) | 1 (16.7) | 2 (50) | 10 (33.3) | 3 (15.8) | 31 (24.8) |  |
|  | Never | 2 (66.7) | 1 (11.1) | 6 (28.6) | 5 (27.8) | 4 (26.7) | 4 (66.7) | 1 (25) | 11 (36.7) | 2 (10.5) | 36 (28.8) |  |
| Dieticians | Routinely | 0 (0) | 0 (0) | 0 (0) | 1 (6.3) | 2 (13.3) | 0 (0) | 0 (0) | 0 (0) | 0 (0) | 3 (2.5) | **0.039** |
|  | Often | 0 (0) | 2 (25) | 4 (19) | 0 (0) | 0 (0) | 0 (0) | 0 (0) | 0 (0) | 1 (5.6) | 7 (5.8) |  |
|  | Sometimes | 0 (0) | 3 (37.5) | 3 (14.3) | 6 (37.5) | 3 (20) | 0 (0) | 0 (0) | 7 (24.1) | 5 (27.8) | 27 (22.5) |  |
|  | Rarely | 1 (33.3) | 1 (12.5) | 5 (23.8) | 3 (18.8) | 5 (33.3) | 1 (16.7) | 2 (50) | 5 (17.2) | 9 (50) | 32 (26.7) |  |
|  | Never | 2 (66.7) | 2 (25) | 9 (42.9) | 6 (37.5) | 5 (33.3) | 5 (83.3) | 2 (50) | 17 (58.6) | 3 (16.7) | 51 (42.5) |  |
| Speech and language pathology | Routinely | 0 (0) | 3 (33.3) | 0 (0) | 2 (11.8) | 0 (0) | 0 (0) | 0 (0) | 0 (0) | 0 (0) | 5 (4.2) | **0.004** |
|  | Often | 0 (0) | 2 (22.2) | 1 (4.8) | 1 (5.9) | 0 (0) | 0 (0) | 0 (0) | 0 (0) | 2 (11.1) | 6 (5) |  |
|  | Sometimes | 0 (0) | 1 (11.1) | 6 (28.6) | 3 (17.6) | 4 (28.6) | 0 (0) | 0 (0) | 6 (21.4) | 5 (27.8) | 25 (20.8) |  |
|  | Rarely | 1 (33.3) | 0 (0) | 3 (14.3) | 1 (5.9) | 4 (28.6) | 1 (16.7) | 2 (50) | 5 (17.9) | 8 (44.4) | 25 (20.8) |  |
|  | Never | 2 (66.7) | 3 (33.3) | 11 (52.4) | 10 (58.8) | 6 (42.9) | 5 (83.3) | 2 (50) | 17 (60.7) | 3 (16.7) | 59 (49.2) |  |
| **Frequency of Interdisciplinary Collaboration in the Management of Syndromic Children with Olfactory Dysfunction** | | | | | | | | | | | | |
| **Specialty** | **Categories** | **Africa** | **Asia** | **Australia** | **Europe** | **Middle East** | **North America** | **South America** | **UK** | **USA** | **Total** | **Chi-square p-value** |
| Paediatric neurology | Routinely | 0 (0) | 8 (72.7) | 0 (0) | 4 (22.2) | 3 (20) | 1 (14.3) | 0 (0) | 1 (3.2) | 3 (15.8) | 20 (15.5) | **<.001** |
|  | Often | 0 (0) | 2 (18.2) | 7 (33.) | 7 (38.9) | 4 (26.7) | 1 (14.3) | 0 (0) | 1 (3.2) | 4 (21.1) | 26 (20.2) |  |
|  | Sometimes | 1 (33.3) | 0 (0) | 7 (33.3) | 4 (22.2) | 4 (26.7) | 1 (14.4) | 0 (0) | 10 (32.3) | 5 (26.3) | 32 (24.8) |  |
|  | Rarely | 0 (0) | 0 (0) | 4 (19) | 0 (0) | 3 (20) | 1 (14.3) | 3 (75) | 13 (41.9) | 6 (31.6) | 30 (23.3) |  |
|  | Never | 2 (66.7) | 1 (9.1) | 3 (14.3) | 3 (16.7) | 1 (6.7) | 3 (42.9) | 1 (25) | 6 (19.4) | 1 (5.3) | 21 (16.3) |  |
| Paediatric endocrinology | Routinely | 0 (0) | 5 (50) | 2 (9.5) | 3 (16.7) | 2 (13.3) | 2 (28.6) | 1 (25) | 4 (12.9) | 2 (11.1) | 21 (16.5) | 0.205 |
|  | Often | 0 (0) | 2(20) | 4 (19) | 5 (27.8) | 3 (20) | 1 (14.3) | 0 (0) | 2 (6.5) | 6 (33.3) | 23 (18.1) |  |
|  | Sometimes | 1 (33.3) | 2 (20) | 9 (42.9) | 6 (33.3) | 5 (3.33) | 2 (28.6) | 0 (0) | 9 (29) | 6 (33.3) | 40 (31.5) |  |
|  | Rarely | 0 (0) | 0 (0) | 4 (19) | 1 (5.6) | 2 (13.3) | 0 (0) | 2 (50) | 10 (32.3) | 3 (16.7) | 22 (17.3) |  |
|  | Never | 2 (66.7) | 1 (10) | 2 (9.5) | 3 (16.7) | 3 (20) | 2 (28.6) | 1 (25) | 6 (19.4) | 1 (5.6) | 21 (16.5) |  |
| Neuropsychology | Routinely | 0 (0) | 1 (12.5) | 0 (0) | 1 (6.3) | 1 (7.1) | 0 (0) | 0 (0) | 0 (0) | 0 (0) | 3 (2.6) | 0.349 |
|  | Often | 0 (0) | 1 (12.5) | 1 (5.3) | 2 (12.5) | 3 (21.4) | 0 (0) | 0 (0) | 0 (0) | 2 (11.8) | 9 (7.9) |  |
|  | Sometimes | 0 (0) | 3 (37.5) | 4 (21.1) | 4 (25) | 3 (21.4) | 0 (0) | 0 (0) | 3 (10.7) | 5 (29.4) | 22 (19.3) |  |
|  | Rarely | 1 (33.3) | 1 (12.5) | 4 (21.1) | 2 (12.5) | 3 (21.4) | 1 (20) | 2 (50) | 6 (21.4) | 7 (41.2) | 27 (23.7) |  |
|  | Never | 2 (66.7) | 2 (25) | 10 (52.6) | 7 (43.8) | 4 (28.6) | 4 (80) | 2 (50) | 19 (67.9) | 3 (17.6) | 53 (46.5) |  |
| Clinical genetics | Routinely | 0 (0) | 4 (40) | 1 (5) | 4 (22.2) | 3 (20) | 2 (28.6) | 0 (0) | 4 (13.3) | 9 (47.4) 2 (10.5) | 27 (21.4) | **0.03** |
|  | Often | 0 (0) | 4 (40) | 6 (30) | 5 (27.8) | 2 (13.3) | 0 (0) | 1 (25) | 1 (3.3) | 3 (15.8) | 21 (16.7) |  |
|  | Sometimes | 0 (0) | 1 (10) | 5 (25) | 6 (33.3) | 3 (20) | 1 (14.3) | 0 (0) | 6 (20) | 4 (21.1) | 25 (19.8) |  |
|  | Rarely | 1 (33.3) | 0 (0) | 2 (10) | 0 (0) | 3 (20) | 1 (14.3) | 2 (50) | 8 (26.7) | 1 (5.3) | 21 (16.7) |  |
|  | Never | 2 (66.7) | 1 (10) | 6 (30) | 3 (16.7) | 4 (26.7) | 3 (42.9) | 1 (25) | 11 (36.7) |  | 32 (25.4) |  |
| Dieticians | Routinely | 0 (0) | 0 (0) | 0 (0) | 0 (0) | 1 (6.7) | 0 (0) | 0 (0) | 1 (3.4) | 0 (0) | 2 (1.7) | **0.039** |
|  | Often | 0 (0) | 2 (25) | 3 (15) | 1 (5.9) | 0 (0) | 0 (0) | 0 (0) | 0 (0) | 2 (11.1) | 8 (6.7) |  |
|  | Sometimes | 0 (0) | 4 (50) | 2 (10) | 6 (35.3) | 5 (33.3) | 0 (0) | 0 (0) | 4 (13.8) | 3 (16.7) | 24 (20) |  |
|  | Rarely | 1 (33.3) | 0 (0) | 6 (30) | 2 (11.8) | 3 (20) | 1 (16.7) | 2 (50) | 7 (24.1) | 11 (61.1) | 33 (27.5) |  |
|  | Never | 2 (66.7) | 2 (25) | 9 (45) | 8 (47.1) | 6 (40) | 5 (83.3) | 2 (50) | 17 (58.6) | 2 (11.1) | 53 (44.2) |  |
| Speech and language pathology | Routinely | 0 (0) | 3 (37.5) | 0 (0) | 1 (5.9) | 1 (7.1) | 1 (14.3) | 0 (0) | 0 (0) | 1 (5.6) | 7 (5.8) | **0.002** |
|  | Often | 0 (0) | 2 (25) | 1 (5) | 1 (5.9) | 0 (0) | 0 (0) | 0 (0) | 0 (0) | 1 (5.6) | 5 (4.2) |  |
|  | Sometimes | 0 (0) | 2 (25) | 4 (20) | 6 (35.3) | 4 (28.6) | 0 (0) | 0 (0) | 6 (20.7) | 5 (27.8) | 27 (22.5) |  |
|  | Rarely | 1 (33.3) | 0 (0) | 5 (25) | 1 (5.9) | 2 (14.3) | 1 (14.3) | 2 (50) | 5 (17.2) | 10 (55.6) | 27 (22.5( |  |
|  | Never | 2 (66.7) | 1 (12.5) | 10 (50) | 8 (47.1) | 7 (50) | 5 (71.4) | 2 (50) | 18 (62.1) | 1 (5.6) | 54 (45) |  |
| **Perceived Barriers to Multidisciplinary Collaboration in the Management of Pediatric Olfactory Dysfunction** | | | | | | | | | | | | |
|  | **Category** | **Africa** | **Asia** | **Australia** | **Europe** | **Middle East** | **North America** | **South America** | **UK** | **USA** | **Total** | **Chi-square p-value** |
|  |  | N (%) | N (%) | N (%) | N (%) | N (%) | N (%) | N (%) | N (%) | N (%) | N (%) |  |
|  | Fragmented healthcare systems | 1 (20) | 3 (18.8) | 3 (11.1) | 5 (21.7) | 4 (22.2) | 4 (40) | 2 (28.6) | 11 (28.2) | 5 (22.7) | 38 (22.8) | 0.784 |
|  | Lack of shared care pathways | 0 (0) | 4 (25) | 10 (37) | 6 (26.1) | 5 (27.8) | 7 (70) | 2 (28.6) | 19 (48.7) | 5 (22.7) | 58 (34.7) | 0.061 |
|  | Lack of referral guidelines | 1 (20) | 9 (56.3) | 13 (48.1) | 11 (47.8) | 5 (27.8) | 5 (50) | 3 (42.9) | 18 (46.2) | 5 (22.7) | 70 (41.9) | 0.39 |
|  | Funding limitations (hospital/institution) | 0 (0) | 1 (6.3) | 9 (33.3) | 1 (4.3) | 0 (0) | 6 (60) | 1 (14.3) | 13 (33.3) | 0 (0) | 31 (18.6) | **<.001** |
|  | Funding limitations (patient/insurance reimbursement) | 1 (20) | 2 (12.5) | 2 (7.4) | 1 (4.3) | 1 (5.6) | 2 (20) | 1 (14.3) | 1 (2.6) | 2 (9.1) | 13 (7.8) | 0.64 |
|  | None – I routinely treat children with OD in collaboration with other specialties | 0 (0) | 1 (6.3) | 3 (11.1) | 5 (21.7) | 2 (11.1) | 0 (0) | 0 (0) | 2 (5.1) | 4 (18.2) | 17 (10.2) | 0.36 |

Supplementary Table 4. Clinician-reported perspectives on multidisciplinary collaboration in the management of pediatric olfactory dysfunction (OD). This includes: (1) frequency of collaboration with various specialties for non-syndromic OD, (2) frequency of collaboration for syndromic OD, and (3) perceived barriers to multidisciplinary collaboration in clinical practice.
